# Eicosapentaenoic acid and Arachidonic acid Protection Against Left Ventricle Pathology: the Multi-Ethnic Study of Atherosclerosis

**DOI:** 10.1101/2024.06.05.24308494

**Authors:** Gregory C. Shearer, Robert C. Block, Shue Huang, Linxi Liu, David M. Herrington, Michael Y. Tsai, Nathan Tintle, Timothy D. O’Connell

**Author notes:** **Corresponding Authors** Gregory C Shearer, PhD Department of Nutritional Sciences 110 Chandlee Laboratory University Park, PA 16802, Timothy D. O’Connell, PhD, Department of Integrative Biology and Physiology University of Minnesota School of Medicine, 3-141 CCRB, 2231 6th Street SE Minneapolis, MN 55414. **Disclosures:** DMH, TDO, and GCS received honoraria for training and advice from Amarin Pharmaceuticals. **Trial Registration** Clinicaltrials.gov: NCT00005487.

## Abstract

**Background:** We have shown that ω3 polyunsaturated fatty acids (PUFAs) reduce risk for heart failure, regardless of ejection fraction status. Ventricular remodeling and reduced ventricular performance precede overt hear failure, however there is little insight into how PUFAs contribute to maladaptive signaling over time. PUFAs are agonists for regulatory activity at g-protein coupled receptors such as Ffar4, and downstream as substrates for monooxygenases (e.g lipoxygenase, cytochrome p450, or cyclooxygenase (COX)) which mediate intracellular adaptive signaling.

**Methods:** Plasma phospholipid PUFA abundance at Exam 1 as mass percent EPA, DHA, and arachidonic acid (AA) from the Multi-Ethnic Study of Atherosclerosis (MESA) were evaluated using pathway modeling to determine the association with time-dependent changes in left ventricular (LV) mass (LVM), end-diastolic LV volume (EDV), and end-systolic volume (ESV) measured by cardiac MRI at Exams 1 and 5. Ejection fraction (EF) and mass:volume (MV) were calculated posteriorly from the first three.

**Results:** 2,877 subjects had available MRI data. Participants with low AA and EPA had accelerated age-dependent declines in LVM. Males with low AA and EPA also had accelerated declines in EDV, but among females there was no PUFA association with EDV declines and exam 5 EDV status was positively associated with AA. Both sexes had nearly the same positive association of AA with changes in ESV.

**Conclusion:** Plasma phospholipid AA and EPA are prospectively associated with indices of heart remodeling, including ventricular remodeling and performance. Combined AA and EPA scarcity was associated with the most accelerated age-related changes and exam 5 status, while the greatest benefits were found among participants with both PUFAs. This suggests that both PUFAs are required for optimal slowing of age-related declines in ventricular function.

## INTRODUCTION

Left ventricular remodeling is a multi-factorial, time-dependent response to aging. In the Multi-Ethnic Study of Atherosclerosis, participants undergoing age-related remodeling replicate a concentric remodeling pattern^1^. This remodeling is accelerated among participants at elevated risk for cardiovascular disease defined similarly to the Framingham risk score. Increasing age is cross-sectionally associated with smaller end-diastolic volume (EDV), end systolic volume (ESV), and left ventricular mass (LVM) in a manner where smaller EDV corresponds with smaller ESV, which when combined, preserves ejection fraction (EF)^2^. Among MESA participants, the smaller negative association of EDV with LVM, was such that mass:volume ratio (M:V) increased with age. In a follow up, LVM increased among male participants over ten years, but did not change among females; EDV declined for both males and females, but the decline was larger among females^3^. ESV declined substantially for females, enough to prevent a decline in EF at the cost of an approximately 10% decrease in SV. In contrast, ESV did not decline for males, which translated to a decline in EF% and an 11% decline in SV, nearly identical to females. In summary, in the MESA cohort, age-related LV remodeling and functional decline were sex dependent but resembled concentric remodeling and were characterized by increasing LVM and moderate declines in EDV among males, but among females it is primarily driven by declining EDV. In both cases, a declining ESV preserved EF^3^.

There is strong clinical interest in understanding the role of PUFAs, particularly ω3-PUFAs (eicosapentaenoic acid, EPA; docosahexaenoic acid, DHA), in cardiac remodelling^4, 5^. In coronary heart disease (CHD), low-dose trials with ω3-PUFAs have produced mixed results with some trials indicating a significant benefit^6^, while other trials indicate no benefit^7^. In heart failure (HF), one older low-dose trial with ω3-PUFAs, Gruppo Italiano per lo Studio della Sopravvivenza nell’Infarto Miocardico-Heart Failure (GISSI-HF), found a reduced incidence of HF or death^8^, which has been supported by other studies^9, 10^. More recently, high-dose trials with ω3-PUFAs have also produced mixed results. In patients with high triglycerides with residual risk despite statin use, the Reduction of Cardiovascular Events with Icosapent Ethyl-Intervention Trial (REDUCE-IT) demonstrated that high-dose EPA (icosapent ethyl, a purified, non-oxidized form of EPA) reduced major adverse coronary events (MACE) by 25%. Conversely, the Statin Residual Risk Reduction with Epanova in High CV Risk Patients with Hypertriglyceridemia (STRENGTH) trial, failed to demonstrate a benefit of high-dose EPA/DHA (carboxylic acids) in a similar patient population.

Explanations for the mixed results in ω3-PUFA clinical trials have understandably focused on the dose and the formulation^5, 11^ or the placebo^12^. However, the molecular mechanism of ω3-PUFA bioactivity is intertwined with the mechanisms of ω6-PUFAs (arachidonic acid, AA) and they share common signaling pathways including: 1) altering membrane structure; 2) signaling through free fatty acid receptors (Ffar) or peroxisome proliferator-activated receptor (Ppars); and/or 3) production of PUFA-derived oxylipins. In the heart, PUFAs can modify remodeling by direct activation of a Ffar4-cytosolic phospholipase A2α (cPLA2α) signaling pathway, which attenuates pathologic remodeling in pressure overload and cardiometabolic disease^13, 14^. Wherever the effects of AA- and EPA-oxylipins on remodeling differ, unique AA×EPA mixtures could be associated with unique outcomes and analytical approaches should properly consider both.

Pathway models are a subset of structural equations^18^ suited to A) account for the association of ω6- and ω3-PUFAs with baseline LVM, EDV, and ESV; B) account for each outcome simultaneously; C) measure the conditional association of PUFAs with outcomes; and finally D) evaluate total effect of PUFAs by separating the direct effect on time-dependent changes from the indirect effects. The Multi-Ethnic Study of Atherosclerosis (MESA) cohort is a well-characterized, population-based, prospective cohort that includes ethnically diverse adults with several important cardiac metrics relating to plasma fatty acids and left ventricular remodeling, including plasma phospholipid fatty acid content and longitudinal left ventricular metrics using MRI^19–23^, three of which can be expressed as mathematically independent outcomes: LVM, EDV, and ESV. In this study, we test hypotheses (A-D), proposing that an interaction between plasma %AA and %EPA is prospectively associated with changes in left ventricular remodeling and function including LVM, EDV, ESV, calculated ejection fraction, and calculated mass:volume ratio.

## PARTICIPANTS AND METHODS

### Study Participants

MESA is a prospective, population-based study designed to investigate the prevalence, risk factors, and progression of subclinical cardiovascular disease in a multi-ethnic cohort in the United States^19, 25^. Its study design, study population, and methods have been described^25^. Between July 2000 and July 2002, 6814 participants aged 45–84 years were recruited from 6 U.S. communities. Exclusion criteria included a history of clinical cardiovascular disease, pregnancy, and weight >300 pounds (136 kg). The MESA study was approved by institutional review boards at each participating study site and all participants gave informed consent. This study samples the subset of participants randomly selected for inclusion in the cardiac MRI study. We excluded anyone with a prior history of cardiovascular disease or non-physiologic fatty acid abundance^9^. 2,877 participants had MRI imaging at both Exam 1 and Exam 5, which allowed for calculation of time-dependent changes, however an additional 2 participants lacking other data were removed to allow for calculation of fit indices.

### Plasma Fatty Acid Measurements

Fasting blood was drawn, EDTA-anticoagulant tubes were collected and processed at the first (baseline) study exam using a standardized protocol as previously described^26^. Briefly, liquid extraction with chloroform/methanol was followed by silica TLC and methylation of the phospholipid band for quantitation by GC/FID. Fatty acid data are reported as percent of total fatty acid peak areas in plasma phospholipids.

### MRI Left Ventricle Measurements

The MESA study used cardiac MRI with 1.5-T magnets to determine left ventricle mass and volumes as previously described.^27^ Briefly, a stack of short-axis images covering the entire left ventricle was acquired with the endocardial and epicardial myocardial borders contoured using a semiautomated method (MASS 4.2, Medis, Leiden, the Netherlands). LV end-diastolic volume and LV end-systolic volume were calculated using Simpson’s rule (the summation of areas on each separate slice multiplied by the sum of slice thickness and image gap). LV mass was determined by the sum of the myocardial area (the difference between endocardial and epicardial contour) times slice thickness plus image gap in the end-diastolic phase multiplied by the specific gravity of myocardium (1.05 g/mL). LV stroke volume was calculated as the difference between LV end-diastolic volume and LV end-systolic volume. The interval between MRI was performed at Exams 1 and 5 was about 9.5 yrs, but this was adjusted to exactly 10 years for all participants^28^. This was achieved by calculating Δparameter/year×10 years.

### Pathway Model

We employed the pathway model approach to simultaneously evaluate the effects of PUFAs, using all of the independent outcomes: left ventricular mass (LVM), end diastolic volume (EDV), and end systolic volume (ESV). Two additional clinically useful outcomes, ejection fraction (EF) and mass:volume ratio (M:V), were calculated posteriorly from the model. to evaluate all three primary outcomes, LVM, EDV, and ESV simultaneously. This approach simultaneously minimizes error across all three outcomes and is useful for causal inferences because it can independently attribute effects to the direct effect of PUFAs on outcomes, or indirectly via their association with Exam 1 values. Therefore, we evaluated how AA and EPA are related to each outcome, and whether the effect of one is dependent on the other.

At baseline, both %AA and %EPA are established from diet, lifestyle, and individual genetics; thus, their abundance *a)* is associated with their historical action on ventricular structure and function, and *b)* has already affected the trajectory of ventricular structure and functional time-dependent changes. This is accounted for by the moderated indirect effect of %AA and %EPA on exam 5 outcome via their association with baseline LVM, EDV, and ESV. The indirect effects occur via the effect of %AA and %EPA on Exam 5 outcomes via Exam 1 (Figure 3, purple pathway). We modeled the moderated direct effects of EPA and AA on 10-year changes in our outcome variables; LVM, EDV, and ESV. For model fit %AA and %EPA were Johnson Su transformed, necessary due to the high skew of %EPA; the graphs presented are inverse transformed. LVM, EDV, and ESV are indexed, which is simply a normalized value, as described by Liao *et al* for the MESA cohort^28^. Log-transformation was sufficient to improve normality for LVM and this was chosen since it preserves a more straightforward, proportional interpretation. EDV and ESV and were normally distributed. Males and females were analyzed as separate groups within the model; parameters were shared by default and allowed to be different when required for model fit.

Model fit was achieved using the AMOS software’s *modification index* to identify candidate parameter constraints that could be freed between sexes. This was employed in an iterative forward-selection process, comparing the adjusted (freed constraints) and unadjusted models using the *nested model comparisons* module that reports the difference in model dF, the p-value associated with the Χ^2^-test, and the difference in Normed Fit Index (NFI), as well as the difference in the Incremental Fit Index (IF), Relative Fit Index (RFI), and Tucker-Lewis Index. Where the model difference was marginal, preference was given to parsimonious models with lower Bayesian Information Criterion (BIC). EF and M:V were calculated posteriorly from the model using the *calculate estimands* feature of AMOS, using bootstrapping (×10,000) to calculate the percentile confidence interval.

### Statistical Methods

Descriptive statistics were used to compare the baseline characteristics of participants using appropriate parametric or non-parametric tests for continuous variables, and the χ2 test for categorical variables. For comparison to the pathway models, regression models were fit using the maximum likelihood approach using SAS 9.4. For development of the pathway model, we used the AMOS 26.0 extension of SPSS v26. JMP 15.0, and GraphPad Prism 7.01 were also used.

## RESULTS

**Participant flow** is outlined in **Figure 1**. Baseline characteristics of all 2,877 participants having an MRI at Exam 1 who returned for a follow up MRI at Exam 5 are described in **Table 1**. The proportion of White (42%), Chinese-American (13%), Black (24%), and Hispanic (21%) participants were nearly the same proportion as all enrolled MESA study participants. Subsequently, two of the 2,877 patients were excluded for having an incomplete data set (**Figure 1**), resulting in a total of 2,875 patients in our analysis.

**Figure 1:**
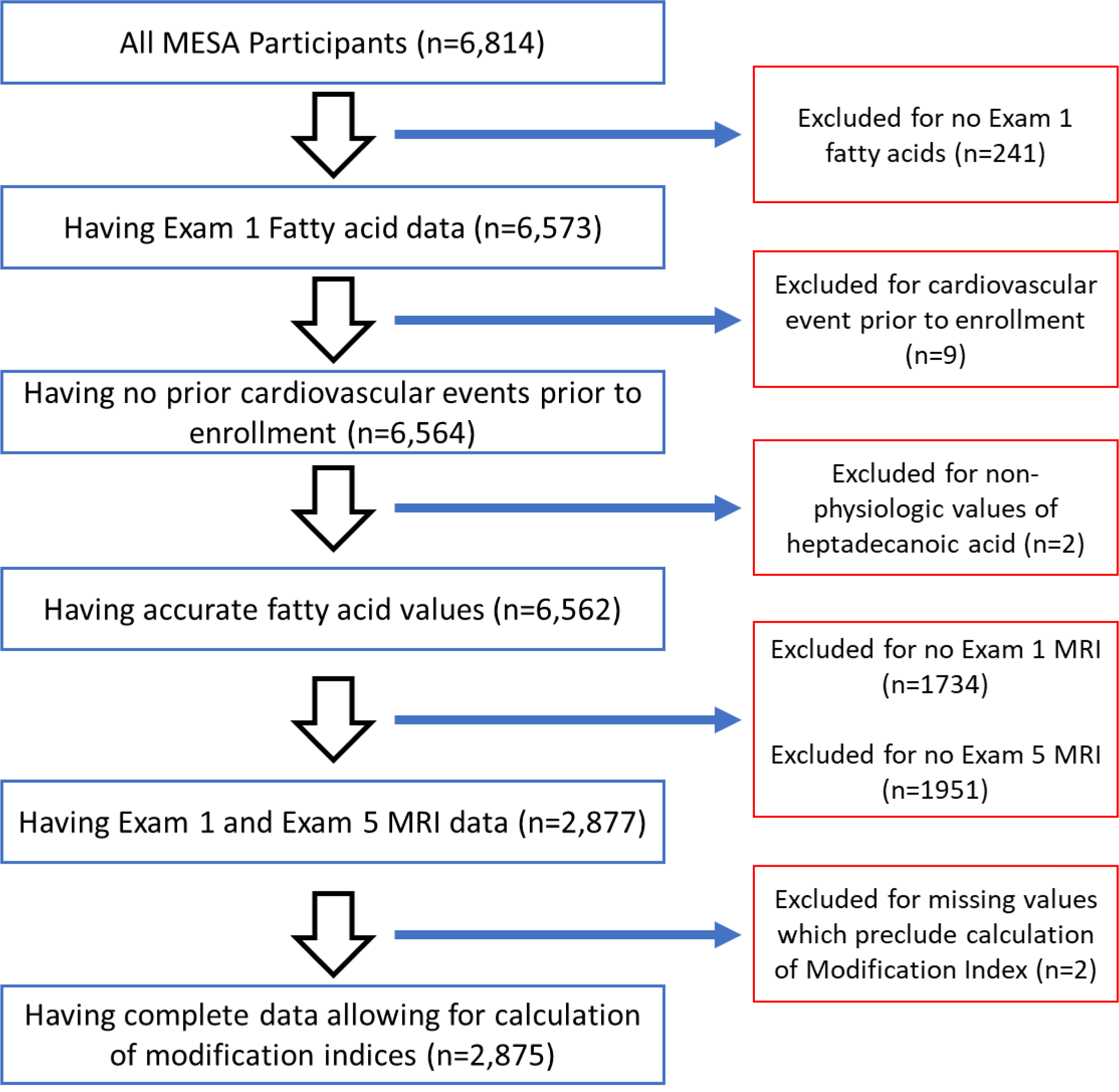
Participant Flow.

**Table 1.** Baseline Characteristics at Exam 1 (N=2877)

| **Variables Mean**<br>**(SD) or N (%)** | **All**<br>**Participants**<br>**(N=2877)** | **White**<br>**(n=1217)** | **Chinese**<br>**(n=369)** | **Black**<br>**(n=701)** | **Hispanic**<br>**(n=590)** | **P-value<sup>A</sup>** |
| --- | --- | --- | --- | --- | --- | --- |
| **Age** |  |  |  |  |  |  |
| Years | 60 (9) | 60 (9) | 58 (9) | 60 (9) | 59 (9) | 0.005 |
| **BMI** |  |  |  |  |  |  |
| (kg/m<sup>2</sup>) | 28 (5) | 27 (5) | 24 (3) | 30 (5) | 29 (4) | <0.0001 |
| **LDL Cholesterol** |  |  |  |  |  |  |
| (mg/dL) | 117 (31) | 116 (30) | 115 (28) | 117 (34) | 121 (32) | 0.02 |
| **Non-HDL**<br>**Cholesterol** |  |  |  |  |  |  |
| (mg/dL) | 143 (35) | 143 (35) | 143 (32) | 137 (36) | 151 (35) | <0.0001 |
| **HDL Cholesterol** |  |  |  |  |  |  |
| (mg/dL) | 51 (15) | 53 (15) | 49 (13) | 54 (16) | 47 (13) | <0.0001 |
| **Triglycerides** |  |  |  |  |  |  |
| (mg/dL) | 129 (75) | 128 (74) | 145 (86) | 100 (50) | 154 (84) | <0.0001 |
| **eGFR** |  |  |  |  |  |  |
| (mL/min/1.73 m<sup>2</sup>) | 82 (16) | 76 (13) | 84 (16) | 87 (18) | 84 (17) | <0.0001 |
| **SBP** |  |  |  |  |  |  |
| Mean (SD) | 123 (20) | 120 (19) | 121 (20) | 129 (20) | 123 (19) | <0.0001 |
| **DBP** |  |  |  |  |  |  |
| Mean (SD) | 72 (10) | 70 (10) | 73 (11) | 75 (10) | 72 (10) | <0.0001 |
| **Heart Rate** |  |  |  |  |  |  |
| Mean (SD) | 62 (9) | 62 (9) | 63 (8) | 62(9) | 62 (9) | 0.35 |
| **Fasting Glucose** |  |  |  |  |  |  |
| (mg/dL) | 94 (25) | 89 (18) | 95 (22) | 97 (29) | 98 (30) | <0.0001 |
| **%EPA** |  |  |  |  |  |  |
| mean (SD) | 1.0 (0.9) | 1 (0.8) | 1.4 (1.3) | 0.9 (0.8) | 0.7 (0.6) | <0.0001 |
| **%DHA** |  |  |  |  |  |  |
| mean (SD) | 4.1 (1.6) | 3.7 (1.4) | 5.5 (1.6) | 4.5 (1.4) | 3.4 (1.3) | <0.0001 |
| **%EPA+DHA** |  |  |  |  |  |  |
| mean (SD) | 5.1 (2.2) | 4.7 (2.1) | 6.9 (2.6) | 5.4 (2.0) | 4.1 (1.7) | <0.0001 |
| **%AA** |  |  |  |  |  |  |
| mean (SD) | 12.3 (2.7) | 11.9 (2.4) | 10.8 (2.2) | 14 (2.5) | 11.8 (2.6) | <0.0001 |
| **Gender** |  |  |  |  |  |  |
| Female (%) | 1511 (53) | 645 (53) | 190 (51) | 401 (57) | 275 (47) | 0.002 |
| Male (%) | 1366 (47) | 572 (47) | 179 (49) | 300 (43) | 315 (53) |  |
| **Diabetes mellitus** |  |  |  |  |  |  |
| Normal % | 2289 (80) | 1065 (88) | 274 (74) | 517 (74) | 433 (73) | <0.0001 |
| Impaired Fasting<br>Glucose % | 336 (12) | 96 (8) | 61 (17) | 94 (13) | 85 (14) |  |
| Untreated Diabetes<br>% | 55 (2) | 12 (1) | 5 (1) | 20 (3) | 18 (3) |  |
| Treated Diabetes % | 194 (7) | 43 (4) | 27 (7) | 70 (10) | 54 (9) |  |
| **Smoking status** |  |  |  |  |  |  |
| Never % | 2602 (90) | 1036 (85) | 367 (99) | 643 (92) | 556 (94) | <0.0001 |
| Former % | 213 (7) | 153 (13) | 1 (0) | 35 (5) | 24 (4) |  |
| Current % | 51 (2) | 27 (2) | 1 (0) | 15 (2) | 8 (1) |  |
| **Urine** |  |  |  |  |  |  |
| **Microalbuminuria** |  |  |  |  |  |  |
| Normal % | 2696 (94) | 1173 (96) | 341 (92) | 642 (92) | 540 (91) | <0.0001 |
| Microalbuminuria % | 150 (5) | 33 (3) | 25 (7) | 50 (7) | 42 (7) |  |
| Macroalbuminuria % | 19 (1) | 6 (0) | 3 (1) | 5 (1) | 5 (1) |  |
| **ACE inhibitor** |  |  |  |  |  |  |
| No % | 2570 (89) | 1104 (91) | 345 (94) | 594 (85) | 527 (89) | <0.0001 |
| Yes % | 299 (10) | 110 (9) | 24 (7) | 104 (15) | 61 (10) |  |
| **ARBs** |  |  |  |  |  |  |
| No % | 2740 (95) | 1173 (96) | 349 (95) | 651 (93) | 567 (96) | 0.004 |
| Yes % | 129 (4) | 41 (3) | 20 (5) | 47 (7) | 21 (4) |  |
| **Beta Blocker** |  |  |  |  |  |  |
| No % | 2634 (92) | 1126 (93) | 332 (90) | 629 (90) | 547 (93) | 0.07 |
| Yes % | 235 (8) | 88 (7) | 37 (10) | 69 (10) | 41 (7) |  |
| **Loop diuretic** |  |  |  |  |  |  |
| No % | 2845 (99) | 1199 (99) | 368 (100) | 692 (9) | 586 (99) | 0.17 |
| Yes % | 24 (1) | 15 (1) | 1 (0) | 6 (1) | 2 (0) |  |
| **Statin** |  |  |  |  |  |  |
| No % | 2471 (86) | 1017 (84) | 329 (89) | 601 (86) | 524 (89) | 0.005 |
| Yes % | 398 (14) | 197 (16) | 40 (11) | 97 (14) | 64 (11) |  |
| **Any hypolipidemic** |  |  |  |  |  |  |
| No % | 2433 (85) | 999 (82) | 321 (87) | 594 (85) | 519 (88) | 0.005 |
| Yes % | 436 (15) | 215 (18) | 48 (13) | 104 (15) | 69 (12) |  |
| **Oral** |  |  |  |  |  |  |
| **Hypoglycemic Agents** |  |  |  |  |  |  |
| No % | 2699 (94) | 1178 (97) | 342 (93) | 641 (91) | 538 (91) | <0.0001 |
| Yes % | 170 (6) | 36 (3) | 27 (7) | 57 (8) | 50 (8) |  |

**Note:** For continuous variables, the rank sum test was used. For most categorical variables, the chi-square test was used. However, for the loop diuretic and urine albumin variables, the exact test was used since they have cells with expected values smaller than 5. Fasting glucose is calibrated, mean (SD). Triglycerides are (use geomean/95% CI). ARBs are angiotensin receptor blockers
<sup>A</sup> Test for group differences

### AA×EPA combinations in MESA participants

As precursors for lipid signaling, AA and EPA occupy the sn2 position of membrane phospholipids, which means that there is an upper limit to the rate of PUFA incorporation into phospholipids, such that increasing one PUFA leads to replacement of the other. In order to evaluate this prediction, the extant mixtures of %AA and %EPA among the MESA participants in this study are shown in **Figure 2**. This analysis identifies limits to their distribution, demonstrating that %AA and %EPA are not strictly independent. We used the 5-10% density quantile to describe the approximate limits of AA×EPA combinations. Given the biochemistry of PUFA incorporation into phospholipids – PUFAs are only incorporated into the *sn*-2 position – phospholipids do not contain more than 50% PUFAs and the presence of limits is expected. Line A in **Figure 2** represents the limit at which increasing EPA incorporation into the *sn*-2 position begins to replace AA (and *vice versa*). Three additional limits were identified (Lines B, C, and D, **Figure 2**) which are; (B) the limit at which greater AA incorporation requires more EPA, (C) the extreme limit of PUFA deficiency, and (D) the limit at which greater EPA incorporation requires greater AA.

**Figure 2:**
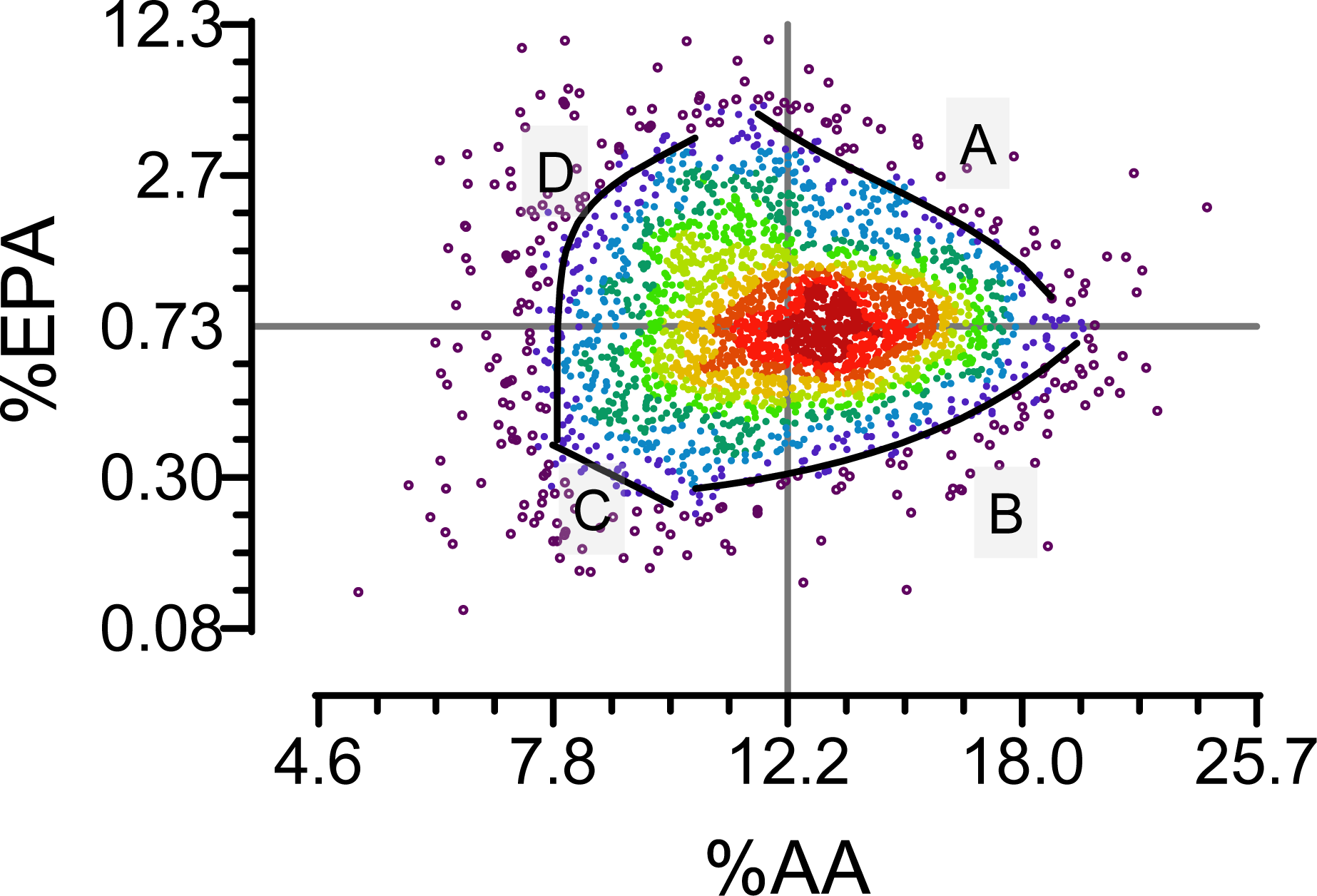
Distributions of AA and EPA as mixture. **AA and EPA – common and rare mixtures** MESA participants are plotted by their %AA (*x*-axis) and %EPA (*y*-axis). Participant density quantile is coded by a color gradient with dark red representing the 95^th^ percentile to purple representing the 5^th^. The highest density combination occurred at 0.69% EPA and 12.6% AA. Boundaries for common mixtures are delineated by participants at the 85th to 90th percentile (bolded) and are evident by bold lines along the top right (A), lower right (B), lower left (C) and left to upper left (D) borders. Note the scale of axes are from the Johnson Su transformation. The limits represented by lines A, B, C, and D have implications as follows: *Line A* implies a maximal limit of PUFA enrichment at which each mass increase in AA is associated with a mass decrease in EPA. *Line B* implies a severity of EPA deficiency which limits maxmal AA content; lower mass enrichment of EPA is associated with a lower maximal AA mass enrichment. *Line C* represents an extreme lower limit of combined AA and EPA deficiency. *Line D* implies a severity of AA deficiency which limits maximal EPA content;

### Univariate and multivariate analyses on PUFAs

We used standard regression models adjusted for covariates in four stages **(Supplemental Tables 1 & 2)**. This allowed us to compare pathway model with a more common approach. In fully adjusted univariate models, we found EPA was positively associated with M:V and EF%, and AA was positively associated with EDV and ESV and negatively associated with M:V **(Supplemental Table 1)**. In fully adjusted multivariate models which include interactions, AA was negatively associated with M:V and combined AA×EPA was associated with LVM and EF% **(Supplemental Table 2)**.

### Final Pathway model

The final model is shown in **Figure 3** with details in Supplemental Tables 3-12. The model fit was good with acceptable *Χ*^2^, CFI, and multiple other indices. Eight sex-independent associations of AA and EPA with ventricular parameters were evident, and a single marginal sex-dependent associations was evident. These parameters were used to derive the AA and EPA associated total changes and final Exam 5 status of each outcome.

**Figure 3:**
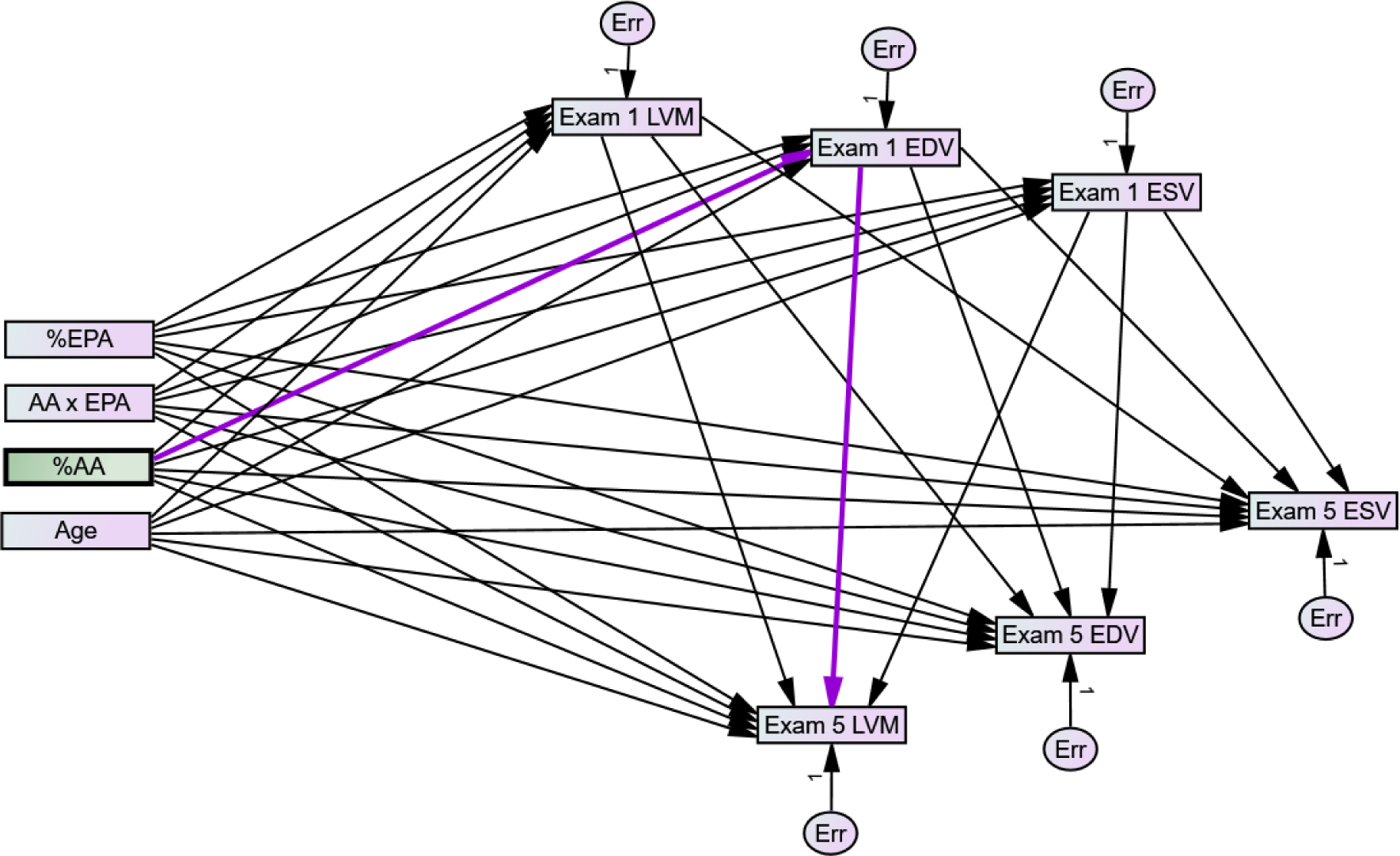
Final pathway model. The final model is depicted using standard format where observed exogenous and endogenous variables are represented by rectangles and unobserved exogenous variables using circles. Paths are represented as linear, single headed arrows and covariances are represented using curved, double-headed arrows. The model allows for indirect effects of AA, EPA, and AA×EPA on Exam 5 (direct arrows to Exam 5 outcomes) or indirect paths by way of Exam 1 outcomes (2 step combination of path to Exam 1 outcome followed by path from Exam 1 to Exam 5 outcome). Highlighted in dark violet is an example of a conditional, indirect effect of AA and EPA on Exam 5 LVM by way of their effect on Exam 1 EDV. Details can be found in **Supplemental Tables**.

### PUFA-dependent effects on LVM at Exam 5

Prior to attribution of PUFA-dependent differences, we calculated the indexed LVM at Exam 1 and Exam 5, which is shown in **Figure 4A & D** (**males & females**). The results indicate a 4.3 index-point decline in LVM in males and a larger 12.6 index-point decline in LVM in females over the 10-year adjusted follow-up period for male and female participants having average %AA and %EPA. The heatmaps in **Figures 4B & E** (**males & females**) show how PUFA-dependent changes in LVM from Exam 1 to 5 are different from the reference change in LVM by AA×EPA combinations, with significant differences outlined. Participants, both male and female, with low %EPA (≤0.74%) had an accelerated decline LVM (green boxes, **Figures 4B & E**), with the greatest acceleration occurring among participants with combined low %AA (≤8.1%) and EPA (≤0.32%). Surprisingly, participants with high %AA, irrespective of EPA status, had a slower decline in LVM over the follow-up (orange boxes, **Figures 4B & E**). The final PUFA-dependent LVM status at Exam 5 strongly corresponded to the PUFA-dependent changes such that in participants with low %AA (≤8.1%) and %EPA (≤0.32%), LVM was smaller than the reference (green boxes, **Figure 4C& F**), whereas in participants with high %AA, irrespective of EPA status, LVM was larger (orange boxes, **Figure 4C & F**).

**Figure 4:**
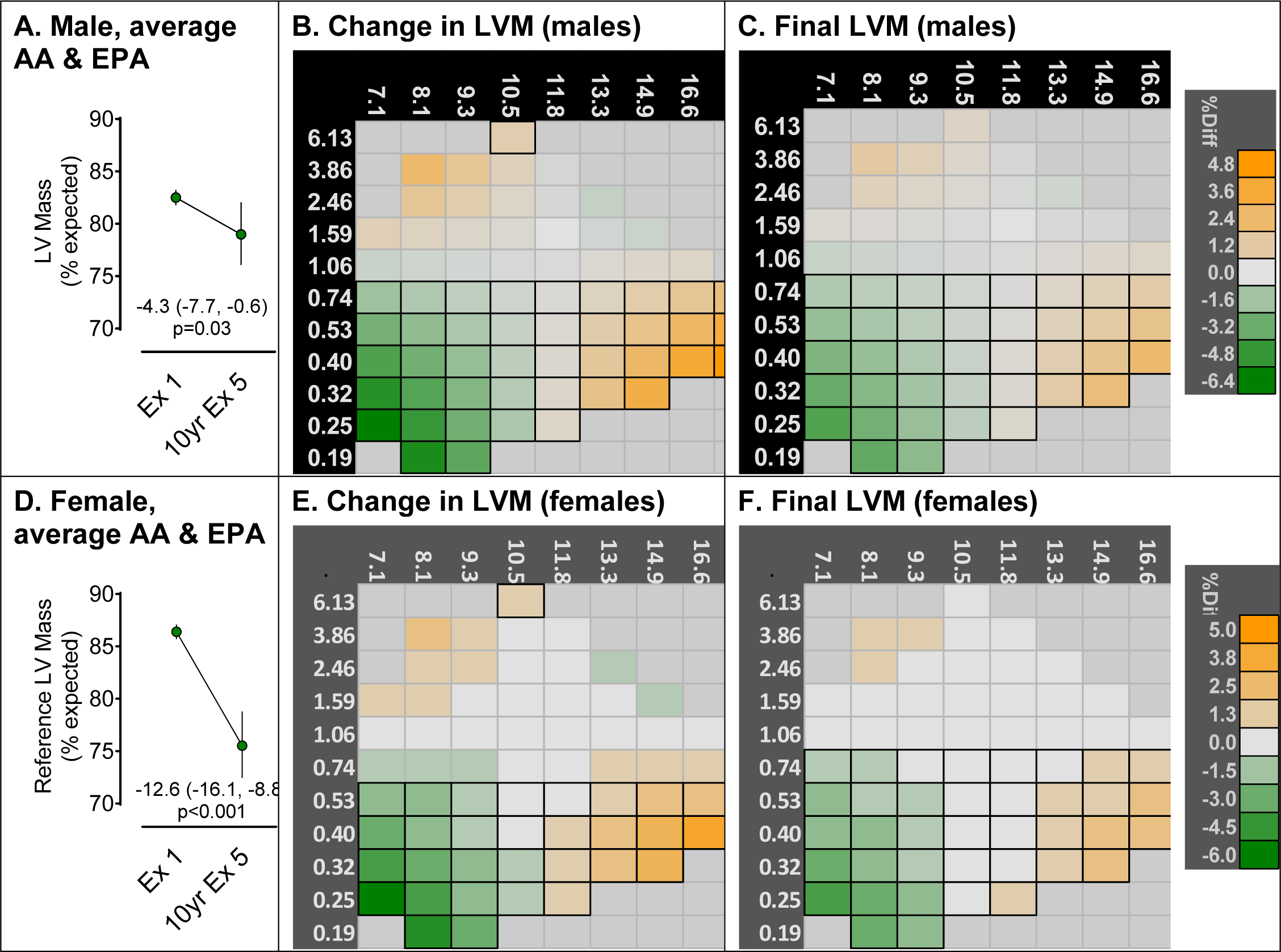
Accelerated loss of LVM when EPA and AA are Both Low. **Heatmap for Exam 5 LVM** by plasma AA (*x*-axis) and EPA (*y*-axis). Panels A and D represent the reference changes in LVM for males and females respectively that are not attributable to PUFAs, either directly or indirectly. These reference values are close to, but less than, the average LVM status reported in Table 2. Panels B and E represent the cumulative 10-year rate of change in LVM in males and females respectively. This is the cumulative change from Exam 1 that is attributable to the direct effect plus the indirect of AA×EPA *via* the associations of AA×EPA with Exam 1 LVM, EDV, and ESV. Panels C and F are the *Final* LVM status at Exam 5 in males and females respectively as it depends on Exam 1 PUFAs. Each square in panels B, C, E, and F represents a unique combination of AA×EPA, and the color corresponds to the proportional difference between the reference Exam 5 LVM. Because accuracy for estimating extreme measures is poor, the heatmap is restricted only to AA×EPA combinations with densities greater than the 5^th^ percentile (See Figure 2). Note the scale of AA and EPA as analyzed is the Johnson Su distribution; the values are converted back to percent total mass for simpler interpretation. Reports of individual PUFA-dependent associations with Exam 1 status and the cumulative indirect effect of Exam 1 LVM, EDV, and ESV on Exam 5 LVM can be found in the supplemental data. Significant differences from reference at p<0.05 is designated by black borders.

### PUFA-dependent effects on EDV at Exam 5

Prior to attribution of PUFA-dependent differences, we calculated the indexed EDV at Exam 1 and Exam 5, which is shown in **Figure 5A & D** (**males & females**). The results indicate a 5.5 index-point decline in EDV in males and a larger 11.2 index-point decline in EDV in females over the 10-year adjusted follow-up period for male and female participants having average %AA and %EPA. The heatmaps in **Figure 5B & E** (**males & females**) show how PUFA-dependent changes in EDV from Exam 1 to 5 are different from the reference change by AA×EPA combinations, with significant differences outlined. Male participants with combined low %AA (≤8.1%) and %EPA (≤0.32%) had an accelerated decline in EDV (green boxes, **Figure 5B**), however the decline in EDV among females was not PUFA-dependent (**Figure 5E**). As a result, the final EDV status at Exam 5 in males with a broad range of combined low %AA and %EPA (%AA≤10.4, %EPA≤0.53) was smaller than the reference EDV (green boxes, **Figure 5C & F; males & females**). In contrast, males with moderate %EPA and high %AA (%AA≥13.3, %EPA≤0.74) had larger EDV than the reference (green boxes, **Figure 5C**), indicating a slower age-dependent decline in EDV among these participants (orange boxes, **Figure 5C**). Among females, the final EDV status corresponded to AA status such that females with low %AA (≤9.5%) and moderate to high %EPA (0.53%≤EPA≤1.59%) had a smaller EDV than reference (green boxes, **Figure 5F**), whereas females with high %AA (%AA≥14.9) had a larger EDV (orange boxes, **Figure 5F**).

**Figure 5:**
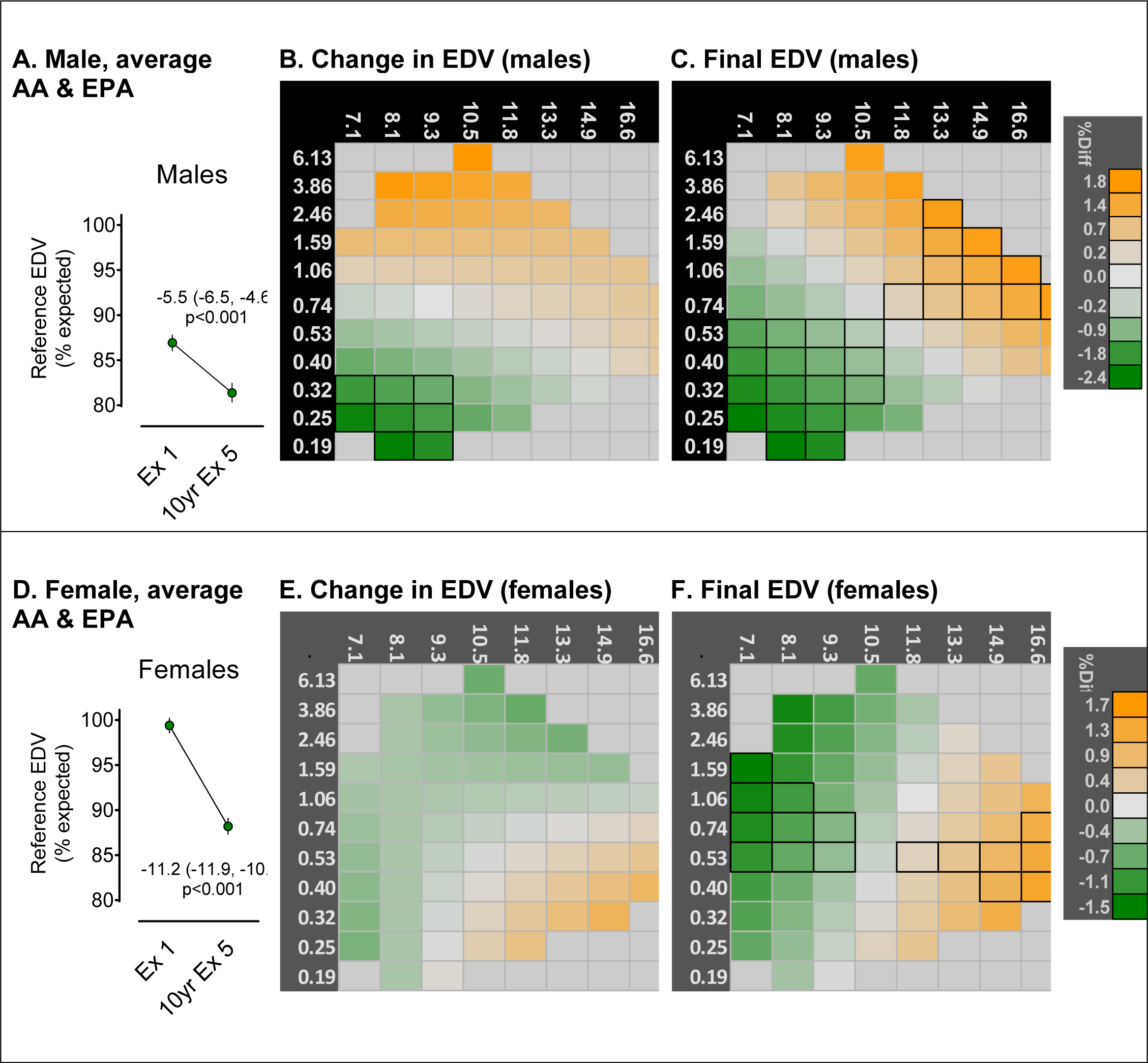
Accelerated loss of EDV when both EPA and AA are Low in Men. **Heatmap for Exam 5 EDV** by plasma AA (*x*-axis) and EPA (*y*-axis). Panels A and D represent the reference changes in EDV for males and females respectively that are not attributable to PUFAs, either directly or indirectly. These reference values are close to, but less than, the average EDV status reported in Table 2. Panels B and E represent the cumulative 10-year rate of change in EDV in males and females respectively. This is the cumulative change from Exam 1 that is attributable to the direct effect plus the indirect of AA×EPA *via* the associations of AA×EPA with Exam 1 LVM, EDV, and ESV. Panels C and F are the *Final* EDV status at Exam 5 in males and females respectively as it depends on Exam 1 PUFAs. Each square in panels B, C, E, and F represents a unique combination of AA×EPA, and the color corresponds to the proportional difference between the reference Exam 5 EDV. Because accuracy for estimating extreme measures is poor, the heatmap is restricted only to AA×EPA combinations with densities greater than the 5^th^ percentile (See Figure 2). Note the scale of AA and EPA as analyzed is the Johnson Su distribution; the values are converted back to percent total mass for simpler interpretation. Reports of individual PUFA-dependent associations with Exam 1 status and the cumulative indirect effect of Exam 1 LVM, EDV, and ESV on Exam 5 EDV can be found in the supplemental data. Significant differences from reference at p<0.05 is designated by black borders.

### PUFA-dependent effects on ESV at Exam 5

Prior to attribution of PUFA-dependent differences, we calculated the indexed ESV at Exam 1 and Exam 5, which is shown in **Figure 6A & D** (**males & females**). The results indicate no decline in ESV in males and a 4.1 index-point decline in ESV in females over the 10-year adjusted follow-up period for male and female participants having average %AA and %EPA. The heatmaps in **Figure 6B & E** (**males & females**) show how PUFA-dependent changes in ESV from Exam 1 to 5 are different from the reference change by AA×EPA concentration, with significant differences outlined. Among both sexes, participants with low %AA (≤10.5%) had accelerated declines in ESV compared to the reference (green boxes, **Figures 6B & E**), however participants with higher %AA (≤10.5%) and moderate EPA (0.32%≤EPA≤0.74%) had slower declines in ESV compared to the reference (orange boxes, **Figures 6B & E**). The final Exam 5 ESV largely corresponded to the 10-year changes in ESV, such that participants with low %AA (≤10.5%) had a smaller ESV compared to reference, whereas participants with higher %AA (≤10.5%) and moderate %EPA (0.32%≤EPA≤0.74%) had a larger ESV (green boxes, **Figures 6C & F**). However while the PUFA-dependent 10-year changes in ESV were not different among sexes, the reference decline among only females resulted in small sex-dependent differences in ESV status at Exam 5 (**Figure 6C & F**).

**Figure 6:**
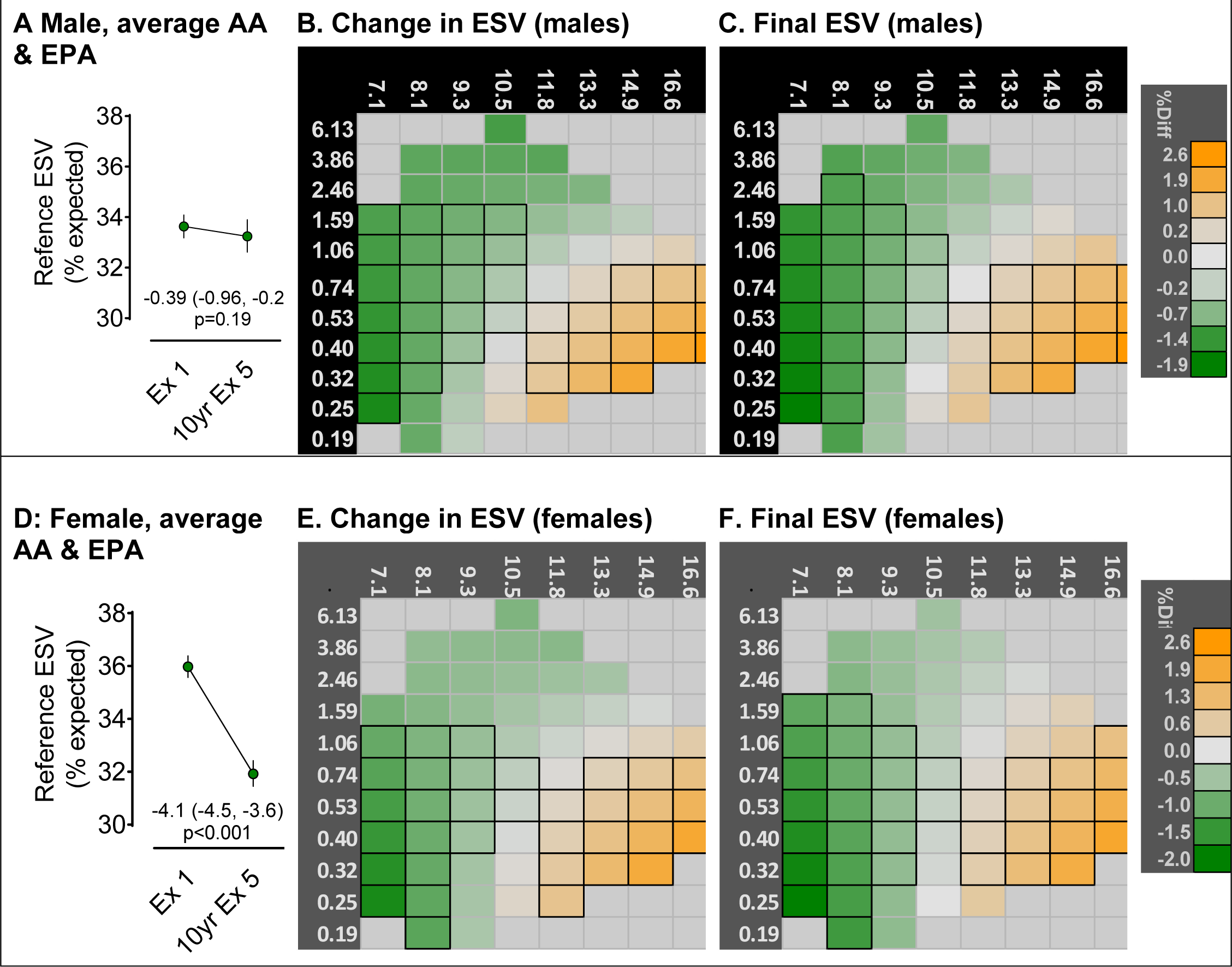
Accelerated decline in ESV among women with EPA is Low and AA Moderate, but delayed decline when EPA is High and AA Moderate in Both Men and Women. **Heatmap for Exam 5 ESV** by plasma AA (*x*-axis) and EPA (*y*-axis). Panels A and D represent the reference changes in ESV for males and females respectively that are not attributable to PUFAs, either directly or indirectly. These reference values are close to, but less than, the average ESV status reported in Table 2. Panels B and E represent the cumulative 10-year rate of change in ESV in males and females respectively. This is the cumulative change from Exam 1 that is attributable to the direct effect plus the indirect of AA×EPA *via* the associations of AA×EPA with Exam 1 LVM, EDV, and ESV. Panels C and F are the *Final* ESV status at Exam 5 in males and females respectively as it depends on Exam 1 PUFAs. Each square in panels B, C, E, and F represents a unique combination of AA×EPA, and the color corresponds to the proportional difference between the reference Exam 5 ESV. Because accuracy for estimating extreme measures is poor, the heatmap is restricted only to AA×EPA combinations with densities greater than the 5^th^ percentile (See Figure 2). Note the scale of AA and EPA as analyzed is the Johnson Su distribution; the values are converted back to percent total mass for simpler interpretation. Reports of individual PUFA-dependent associations with Exam 1 status and the cumulative indirect effect of Exam 1 LVM, EDV, and ESV on Exam 5 ESV can be found in the supplemental data. Significant differences from reference at p<0.05 is designated by black borders.

### PUFA-dependent effects on EF_calc_ at Exam 5

Reference Exam 1 and Exam 5 EF values were calculated from the model as estimands using EDV and ESV as:

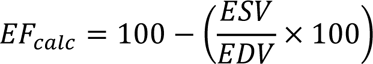

Prior to attribution of PUFA-dependent differences, we determined EF*_calc_* at Exam 1 and Exam 5, which is shown in **Figure 7A & D** (**males & females**). The results indicate a 2.2% decline in EF*_calc_* in males but no change in EF*_calc_* in females over the 10-year adjusted follow-up period for participants having average %AA and %EPA. The heatmaps in **Figure 7B & E** (**males & females**) show how PUFA-dependent changes in EF*_calc_* from Exam 1 to 5 are different from the reference change by AA×EPA combinations, with significant differences outlined. Male participants along a diagonal favoring higher %EPA (≥0.32%) and %low AA (≤13.3%) had slower declines in EF*_calc_* compared to the reference (green boxes, **Figure 7B**), while high %AA and low %EPA had accelerated declines EF*_calc_* (orange boxes, **Figure 7B**), which corresponded to the final EF*_calc_* status (**Figure 7C**). Since no change in EF*_calc_* status was observed among females the panel E changes (**Figure 7E**) represent absolute time-dependent changes in EF*_calc_*. and correspond nearly entirely with final status (**Figure 7F**). PUFA-dependent effects were observed only for female participants with low %EPA (≤1.59%), and were inversely associated with %AA such that females with low %AA (≤10.5%) had increases in EF*_calc_* (orange boxes, **Figure 7F**), however those with higher %AA (≥11.8%) had declines (green boxes, **Figure 7F**).

**Figure 7:**
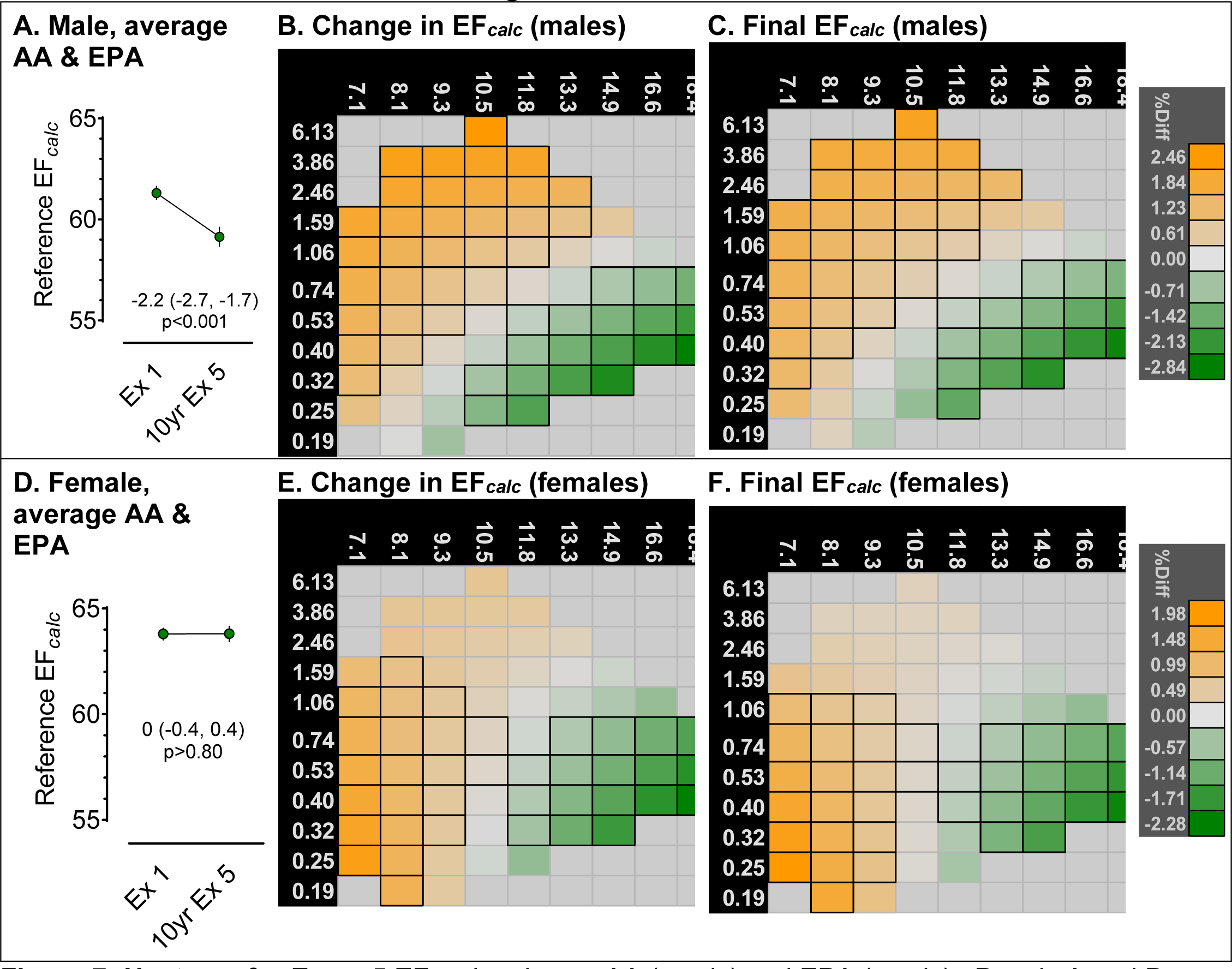
Heatmap for Exam 5 Showing that EF*_calc_* is Reduced when EPA is High and AA Moderate and Increased when AA is High and EPA Low to Moderate. **Heatmap for Exam 5 EF*_calc_*** by plasma AA (*x*-axis) and EPA (*y*-axis). Panels A and D represent the reference changes in EF*_calc_* for males and females respectively that are not attributable to PUFAs, either directly or indirectly. These reference values are close to, but less than, the average EF*_calc_* status reported in Table 2. Panels B and E represent the cumulative 10-year rate of change in EF*_calc_* in males and females respectively. This is the cumulative change from Exam 1 that is attributable to the direct effect plus the indirect of AA×EPA *via* the associations of AA×EPA with Exam 1 LVM, EDV, and ESV. Panels C and F are the *Final* EF*_calc_* status at Exam 5 in males and females respectively as it depends on Exam 1 PUFAs. Each square in panels B, C, E, and F represents a unique combination of AA×EPA, and the color corresponds to the proportional difference between the reference Exam 5 EF*_calc_*. Because accuracy for estimating extreme measures is poor, the heatmap is restricted only to AA×EPA combinations with densities greater than the 5^th^ percentile (See Figure 2). Note the scale of AA and EPA as analyzed is the Johnson Su distribution; the values are converted back to percent total mass for simpler interpretation. Reports of individual PUFA-dependent associations with Exam 1 status and the cumulative indirect effect of Exam 1 LVM, EDV, and ESV on Exam 5 EF*_calc_* can be found in the supplemental data. Significant differences from reference at p<0.05 is designated by black borders.

### PUFA-dependent effects on M:V_calc_ at Exam 5

Reference Exam 1 and Exam 5 EF values were calculated from LVM and EDV as:

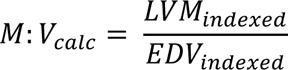

Prior to attribution of PUFA-dependent differences, we determined the M:V*_calc_* at Exam 1 and Exam 5, which is shown in **Figure 8A & D** (**males & females**). The results indicate a 0.241 absolute increase in the M:V ratio in males and a 0.022 absolute increase in the M:V ratio in females over the 10-year adjusted follow-up period for reference male and female participants. The heatmaps in **Figure 8B & E (males & females)** show how PUFA-dependent changes in M:V*_calc_* from Exam 1 to 5 are different from the reference change by AA×EPA combinations, with significant differences outlined. Male participants with high %EPA (≥1.6%) and low %AA (≤10.5%) had accelerated increases in M:V*_calc_*, compared to the reference increase (orange boxes, **Figure 8B**), however higher %AA and lower EPA effectively reversed this accelerated increase (green boxes, **Figure 8B**). Still, the final PUFA-dependent M:V*_calc_* status at Exam 5 was not different by PUFAs (**Figure 8C**). Females with PUFA scarcity or AAxEPA abundance had slower increases in M:V*_calc_* compared to the reference (green boxes, **Figure 8E**). The final M:V*_calc_* status at Exam 5 was lower in females with low %AA (≤9.3%) (green boxes, **Figure 8F**).

**Figure 8:**
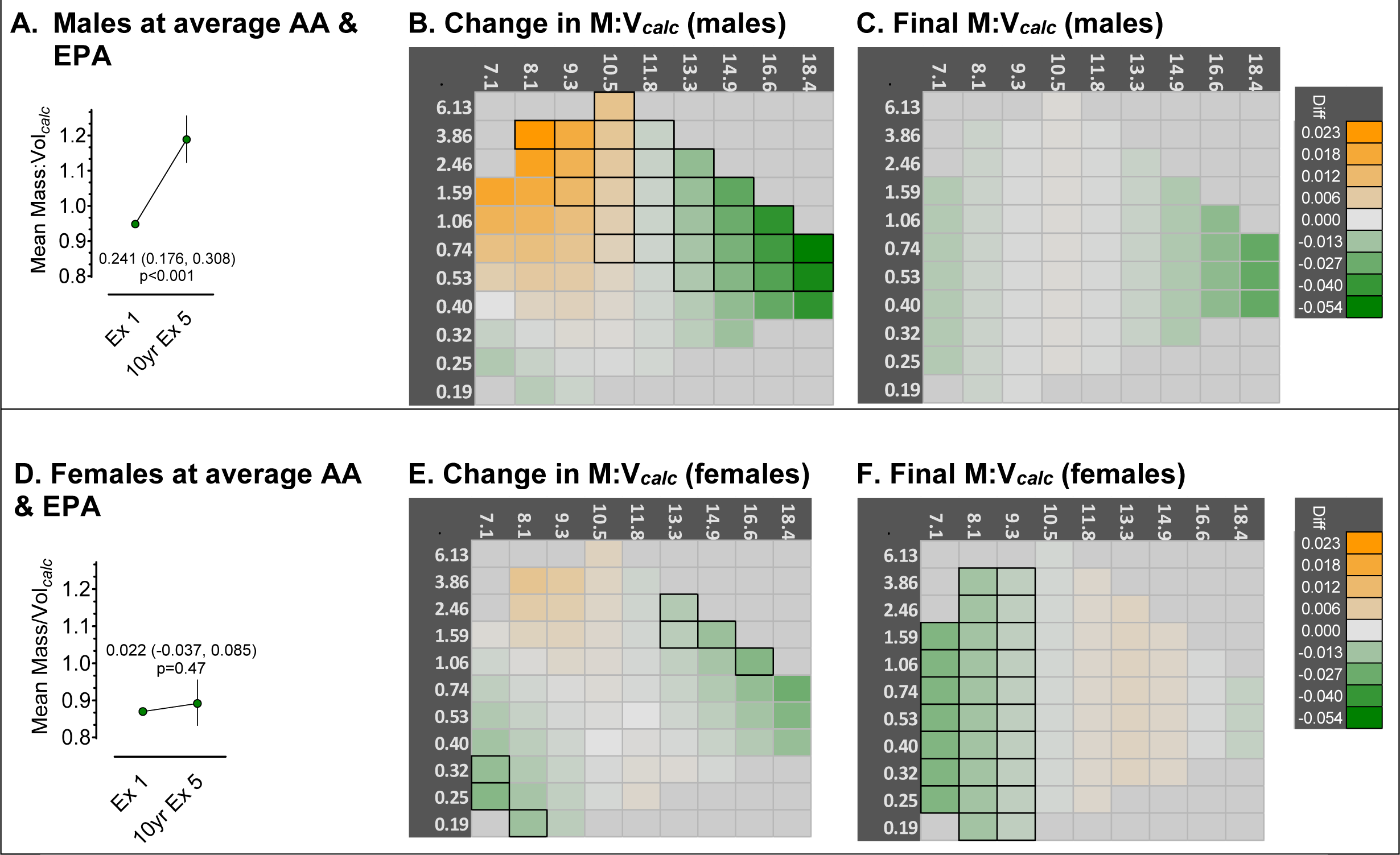
Heatmap for Exam 5 M:V*_calc_*. **Heatmap for Exam 5 M:V*_calc_*** by plasma AA (*x*-axis) and EPA (*y*-axis). Panels A and D represent the reference changes in M:V*_calc_*, males and females respectively, that are not attributable to PUFAs either directly or indirectly. The reference value is an estimate of the M:V*_calc_* when AA and EPA *z*-scores are zero. Panels B and E represent the cumulative 10-year rate of change in M:V*_calc_* in males and females respectively. This is the cumulative change from Exam 1 that is attributable to the direct effect plus the indirect of AA×EPA *via* the associations of AA×EPA with Exam 1 LVM, EDV, and ESV. Panels C and F are the *Final* M:V*_calc_* status at Exam 5 in males and females respectively as it depends on Exam 1 PUFAs. Each heatmap square in panels B, C, E, and F represents a unique combination of AA×EPA, and the color corresponds to the difference from the reference Exam 5 M:V*_calc_*. Because accuracy for estimating extreme measures is poor, the heatmap is restricted only to AA×EPA combinations with densities greater than the 5^th^ percentile (See Figure 2). Note the scale of AA and EPA as analyzed is the Johnson Su distribution; the values are converted back to percent total mass for simpler interpretation.

## DISCUSSION

In this study, we show the complexity and nuanced associations of time-dependent changes in left ventricular properties with AA and EPA. In each case, the identified associations of EPA with LVM, EDV, and ESV over the 10-year adjusted follow-up period as well as the final Exam 5 status were dependent on how much AA was present and *vice versa*. The broadest finding was a delay in age-related changes in LVM, EDV, and ESV among participants having abundant AA and EPA, but accelerated changes among those lacking AA and EPA. The common narrative that EPA is beneficial was largely confirmed, but was also nuanced and explained in more detail below. We also identified multiple contexts where AA was beneficial or where specific, high abundance AA×EPA mixtures were most beneficial, countering the simple narrative “more EPA is good/more AA is bad”. We were surprised to find that EF*_calc_* was actually increased among PUFA-poor participants, which was the result of a more rapid decline in EDV than in ESV. Counter-intuitively, participants with high EPA and moderate AA also had higher EF*_calc_* values, but this was because of a slower decline in EDV paired with no change in ESV. It seems unlikely that the increased EF*_calc_* in PUFA poor participants represents the same remodeling dynamics as the increased EF*_calc_* among PUFA rich participants. Finally, we propose that the identified interactions of AA and EPA on affecting left ventricular properties have strong concordance with the molecular mechanism of action for PUFA mediated signaling, and hence show promise for understanding their role in remodeling dynamics.

To summarize the associations of PUFAs with time-dependent changes it is useful to 1) characterize them against this backdrop of ventricular aging, 2) consider the relative abundances of AA×EPA mixtures characterized against the backdrop of the Lines from Figure 2: the first group along line C, having combined low AA/EPA; the second along the right side of line B having high AA but low EPA; the third along the high end of line D having high EPA but low AA; and the fourth having the maximum plausible combination of AA×EPA, nearly in the middle of line A.

### Participants having low AA and EPA (line C)

Participants with low AA and EPA showed strong evidence for accelerated aging across each outcome, among both sexes, and irrespective of PUFA-dependent changes or the final PUFA-dependent status. These participants experienced the largest declines in LVM and had the smallest LVMs at Exam 5, consistent with accelerated age-related declines. PUFA-deplete males experienced the largest declines in EDV and had the smallest EDV status at Exam 5, despite beginning from Exam 1 at nearly the mean EDV. Their decline in ESV did not quite preserve overall SV, but either maintained or nearly maintained EF. In contrast to males, the aging pattern among females was characterized by more rapidly declining ESV and consequently SV rather than EDV which counter-intuitively increased EF, an exaggeration of the pattern described by Liu et al^1^.

### High levels of either AA (line B) or EPA (line D) were sufficient to delay age-related changes, but, high combined EPA and AA (line A) had the greatest delay in age-related changes

Loss of LVM was delayed among participants with high AA, whereas high EPA normalized the rate of LVM loss, a benefit for participants with low AA status but an acceleration for those with high AA status. For males, slower total declines in ESV were associated with high AA status. This makes interpretation of total changes in EF status more complex since the total decline in EF among male participants with high AA occurred because of delayed decline in both total EDV and total EDV. However among participants with high EPA, total increased EF was due delayed total declines in EDV paired with accelerated total declines in ESV. PUFAs had no total effect on EDV among females; the primary changes among were driven by age-related total declines in ESV, and so female participants lacking AA had total preservation of EF because their total age-related decline in ESV was accelerated. The Exam 5 status of females was best among participants with high AA status who had delayed loss of LVM, the largest EDV, and the smallest decline in ESV all associated with delayed aging. Ironically, the combined delayed aging pattern meant that this group had a decline in EF. It is unclear whether this delayed aging is also associated with increased risk for HF or other pathology.

In the past, when assessing the relationship between fatty acids and ventricular remodeling, we and others have focused on univariate, monotonic approaches because of their straightforward analyses and interpretation^9, 10^. In the full MESA cohort (N=6,814), greater ω3-PUFAs at Exam 1 were associated with reduced hazards for heart failure, regardless of heart failure type^9^. Similar findings were reported in the Cardiovascular Health Study (CHS), a prospective cohort study from 1992 to 2006 that included 2,735 U.S. adults from 4 US communities without prevalent heart disease^10^. In the CHS, there was an association between plasma phospholipid %EPA and an approximately 50% reduction in incident HF in the highest versus the lowest quartile. Fewer analyses have focused on AA, however a non-linear U-shaped association with acute coronary syndrome suggests a complex relationship where AA is beneficial at optimal, but not extreme, levels^29^. Here, we found multiple ways in which AA is associated with beneficial effects, either independently or conditionally: with delayed age-dependent loss of mass, with larger than average EDV at Exam 5, and with delayed age-related decline in ESV. Likewise, EPA was associated delayed age-dependent changes, depending on the availability of AA. Notably, the pathway model approach was successful in identifying these relationships where multiple linear regression analyses were not.

The basis for the EPA-*good*/AA-*bad* narrative is the molecular utilization of PUFAs as precursors for intracellular lipid signaling (**Figure 9**)^4^. Within this framework, both AA and EPA are required for adaptive signaling. However, oxylipins produced from AA have potent pro-inflammatory activity, whereas oxylipins derived from EPA have either neutral or anti-inflammatory activity. At the molecular level, neither PUFA can be proportionally increased in phospholipids without displacing the other’s availability. This is evident in the limits to achieving combined AA and EPA levels, and we found that few participants achieved combined relative AA×EPA abundances to the upper right of line A. Participants were unable to achieve these higher AA×EPA abundances because PUFAs only occupy the *sn*2 position of phospholipids, and so the maximal PUFA occupancy is 50%. This limit also includes other PUFAs essential for signaling such as the 22-carbon ω3-DHA and ω6- docosapentaenoic acid (DPA), as well as the 18-carbon linoleic acid (LA) and α-linolenic acid (aLA). Line A translates to between 31% and 38% of all PUFAs acylated at the phospholipid *sn*2 position. Hence, higher AA×EPA abundances are not achievable since they are displacing each other and other PUFAs, a dynamic that consequently impairs dependent adaptive signaling. More surprising, when one or both PUFAs were scarce there were also limits. Line D represents a limit at which further EPA incorporation necessitated some additional AA (line B) and line B represents the reverse (D). The last line, Line C represents the lower threshold of combined AA×EPA tolerance, the group most consistently characterized by accelerated aging. Ventricular changes in this group should be considered as potentially driven by PUFA scarcity.

Our findings also have implications for interventions, a particularly confusing field where we have noted seemingly similar trials have yielded conflicting results (positive^6, 8, 30^, negative^7, 31^). Notably, current interventions focus exclusively on ω3-PUFAs, and we can predict well how ω3-PUFA interventions will predict changes in ω3-PUFA availability and tissue enrichment^32^. However, increasing ω3-PUFA intake also results in reduced AA content^33, 34^. Here, we add two more considerations: since outcomes are dependent on both AA and EPA, it is not clear that every AA×EPA combination will benefit from an intervention that increases EPA at the cost of reduced AA; second, the starting AA×EPA combination a participant begins from itself has a remodeling trajectory that will impact the need for an intervention. We recommend that assessment in RCTs in outcomes-based trials assess the baseline and final AA×EPA conditions, and their impact on endpoints.

### Strengths and Limitations

Strengths of this study include: the pathway model approach including cross-sectional and longitudinal data; the broad population studied including African-American (28%), Hispanic (22%), Chinese-American (12%), and Caucasians (38%) from 6 US communities; the use of validated MRI data allow for time comparisons^35–37^; and a high-quality plasma phospholipid fatty acid measurement method^26^. Limitations include the fact that plasma levels of EPA and DHA in this cohort study have a limited dynamic range and may be biased towards AA levels which would overrepresent the harms of AA and the benefits of EPA. Plasma phospholipids are less representative of chronic intake compared to RBC membrane levels. As planned, the pathway model lacks adjustment for covariates. This is because the molecular theory for PUFA action identifies the covariates being mediated by blood pressure, inflammation, blood lipids, etc. We were interested in finding the absolute effect of PUFAs, not the effects of PUFAs that are independent of others. Adjusted models are available in the supplemental data. Our interpretation depends on normal age-related LV remodeling being distinct from pathological remodeling.

In conclusion, age related changes are PUFA-dependent, where the association of one PUFA with remodeling outcomes is dependent on the availability of the other. While we did confirm that increased availability of EPA is associated with counteracting age-related changes in ventricular mass and performance, two counter-narrative findings add nuance which could contribute to more accurate risk and clinical management: first, AA also was associated with counteracting age-related changes; second, in every case PUFA scarcity was associated with accelerated age-related changes. Further, PUFA scarcity was counter-intuitively associated with two common clinically useful tools: EF and M:V in which seemingly beneficial changes occur in participants with PUFA scarcity more likely reflect accelerated aging than benefit.

## Supporting information

Supplemental Results and Figures

## Data Availability

All data produced in the present study are available upon request from the MESA study committee

## Notes

**Sources of Support**: This publication was made possible by Grant Numbers 1 R01 HL130099-01A1 and 1 R01 HL152215-01 from the National Heart, Lung, and Blood Institute (TDO, GCS). This research was also supported by contracts HHSN268201500003I, N01-HC-95159, N01-HC-95160, N01-HC-95161, N01-HC-95162, N01-HC-95163, N01-HC-95164, N01-HC-95165, N01-HC-95166, N01-HC-95167, N01HC-95168, andN01-HC-5169 from the National Heart, Lung, and Blood Institute and by grants UL1-TR-000040, UL1-TR001079, and UL1-TR-001420 from the National Center for Advancing Translational Sciences. The content is solely the responsibility of the authors and does not necessarily represent the official views of the NHLBI or NIH.

### Competing Interest Statement

The authors have declared no competing interest.

### Funding Statement

This publication was made possible by Grant Numbers 1 R01 HL130099-01A1 and 1 R01 HL152215-01 from the National Heart, Lung, and Blood Institute (TDO, GCS). .This research was also supported by contracts HHSN268201500003I, N01-HC-95159, N01-HC-95160, N01-HC-95161, N01-HC-95162, N01-HC-95163, N01-HC-95164, N01-HC-95165, N01-HC-95166, N01-HC-95167, N01HC-95168, andN01-HC-5169 from the National Heart, Lung, and Blood Institute and by grants UL1-TR-000040, UL1-TR001079, and UL1-TR-001420 from the National Center for Advancing Translational Sciences.

### Author Declarations

The IRB committee of The Pennsylvania State University gave ethical approval for this work.

