## Supplemental Results and Figures for "Eicosapentaenoic acid and Arachidonic acid Protection Against Left Ventricle Pathology: the Multi-Ethnic Study of Atherosclerosis"

**Supplemental Table 1: Univariate association of outcomes with EPA, DHA, and AA (regression coefficients Beta, SD and P-value)**

| | **10-year Mass Index (log_g/$m^{2.7}$)** | | | **10-year End Diastolic Volume Index (log_ml/$m^{2.7}$)** | | | **10-year Mass to Volume ratio (log_g/ml)** | | | **10-year Ejection Fraction (log_%)** | | | **10-year Stroke volume Index (log_ml/$m^{2.7}$)** | | | **10-year End systolic Volume Index (log_ml/$m^{2.7}$)** | | |
|---|---|---|---|---|---|---|---|---|---|---|---|---|---|---|---|---|---|---|
| | **Beta** | SD | P-value | **Beta** | SD | P-value | **Beta** | SD | P-value | **Beta** | SD | P-value | **Beta** | SD | P-value | **Beta** | SD | P-value |
| EPA | | | | | | | | | | | | | | | | | | |
| model 1 | -0.00170 | 0.0045 | 0.70 | -0.0057 | 0.0056 | 0.31 | 0.0019 | 0.0056 | 0.73 | **0.0076** | **0.0037** | **0.04** | -0.0015 | 0.0065 | >0.80 | -0.0088 | 0.0082 | 0.28 |
| model 2 | 0.00065 | 0.0045 | 0.88 | -0.0068 | 0.0058 | 0.24 | 0.011 | 0.0058 | 0.07 | **0.0078** | **0.0038** | **0.04** | 3.1E-05 | 0.0069 | >0.80 | -0.012 | 0.0084 | 0.15 |
| model 3 | 0.00065 | 0.0045 | 0.88 | -0.0073 | 0.0058 | 0.21 | **0.014** | **0.0059** | **0.02** | **0.0078** | **0.0038** | **0.04** | 3.1E-05 | 0.0069 | >0.80 | -0.013 | 0.0084 | 0.13 |
| model 4 | 0.00420 | 0.0044 | 0.34 | -0.0100 | 0.0058 | 0.07 | **0.019** | **0.0057** | **0.0008** | **0.0075** | **0.0038** | **0.049** | -0.0033 | 0.0068 | 0.62 | -0.016 | 0.0084 | 0.06 |
| DHA | | | | | | | | | | | | | | | | | | |
| model 1 | 0.0036 | 0.0072 | 0.62 | 0.0140 | 0.0089 | 0.11 | -0.016 | 0.009 | 0.08 | **0.015** | **0.0059** | **0.01** | 0.019 | 0.010 | 0.06 | 0.004 | 0.013 | 0.76 |
| model 2 | -0.0035 | 0.008 | 0.67 | 0.0065 | 0.01 | 0.53 | -0.0091 | 0.01 | 0.38 | 0.01 | 0.0068 | 0.13 | 0.01 | 0.012 | 0.40 | -0.000061 | 0.015 | >0.80 |
| model 3 | -0.0035 | 0.008 | 0.67 | 0.0025 | 0.01 | 0.81 | -0.022 | 0.013 | 0.10 | 0.01 | 0.0068 | 0.13 | 0.01 | 0.012 | 0.40 | -0.0058 | 0.015 | 0.70 |
| model 4 | 0.0045 | 0.0078 | 0.56 | -0.0048 | 0.01 | 0.65 | -0.0051 | 0.013 | 0.69 | 0.0089 | 0.0068 | 0.19 | 0.011 | 0.012 | 0.37 | -0.0098 | 0.015 | 0.52 |
| AA | | | | | | | | | | | | | | | | | | |
| model 1 | 0.02700 | 0.012 | 0.03 | **0.052** | **0.015** | **0.0007** | -0.018 | 0.016 | 0.24 | **-0.028** | **0.010** | **0.007** | 0.028 | 0.018 | 0.13 | **0.0896** | **0.023** | **<0.0001** |
| model 2 | 0.00998 | 0.013 | 0.43 | **0.048** | **0.016** | **0.003** | **-0.0396** | **0.016** | **0.02** | -0.013 | 0.010 | 0.21 | 0.037 | 0.019 | 0.053 | **0.067** | **0.024** | **0.005** |
| model 3 | 0.00998 | 0.013 | 0.43 | **0.048** | **0.016** | **0.003** | -0.027 | 0.017 | 0.11 | -0.013 | 0.010 | 0.21 | 0.037 | 0.019 | 0.053 | **0.067** | **0.024** | **0.005** |
| model 4 | 0.00420 | 0.012 | 0.73 | **0.045** | **0.016** | **0.005** | **-0.038** | **0.016** | **0.02** | -0.021 | 0.011 | 0.05 | 0.033 | 0.019 | 0.09 | **0.065** | **0.024** | **0.006** |

Model 1: adjust for outcome at exam 1

Model 2: model 1+ age, sex, race, study center;

Model 3: model 2+ selected variables from fatty acid cluster leads, including trans-oleic acid, palmitoleic acid, behenic acid, gadoleic acid, arachidonic acid, myristic acid, stearic acid, using stepwise with BIC as model selection criteria;

Model 4: model 3 + selected variables from smoking, BMI, nonhdl, TG, EGFR, systolic blood pressure, diastolic blood pressure, heart rate, ACEI, ARB, beta-blockers, fasting glucose, diabetes, oral hypoglycemic, diuretics, statin, hypolipidemic, albuminuria, using stepwise with BIC as model selection criteria

**Supplemental Table 2 Regression models for association of cardiac remodeling measurements with EPA, DHA and their interaction with AA (regression coefficients Beta, SD and P-value)**

| | | 10-year Mass Index (log_g/m$^{2.7}$) | | | 10-year Volume Index (log_ml/m$^{2.7}$) | | | 10-year Mass to Volume ratio (log_g/ml) | | | 10-year Ejection Fraction (log_%) | | | 10-year Stroke volume Index (log_ml/m$^{2.7}$) | | | 10-year End systolic Volume Index (log_ml/m$^{2.7}$) | | |
|---|---|---|---|---|---|---|---|---|---|---|---|---|---|---|---|---|---|---|---|
| | | **Beta** | SD | P-value | **Beta** | SD | P-value | **Beta** | SD | P-value | **Beta** | SD | P-value | **Beta** | SD | P-value | **Beta** | SD | P-value |
| Model 1 | **EPA** | **-0.12** | **0.044** | **0.007** | -0.06 | 0.055 | 0.28 | -0.037 | 0.056 | 0.51 | **0.08** | **0.037** | **0.02** | 0.026 | 0.065 | 0.69 | **-0.18** | **0.081** | **0.02** |
| | **AA** | **-0.24** | **0.099** | **0.02** | -0.068 | 0.12 | 0.58 | -0.1 | 0.12 | 0.40 | 0.14 | 0.082 | 0.08 | 0.09 | 0.14 | 0.53 | -0.3 | 0.18 | 0.09 |
| | **Interaction** | **-0.053** | **0.02** | **0.008** | -0.024 | 0.025 | 0.32 | -0.017 | 0.025 | 0.49 | **0.04** | **0.016** | **0.04** | 0.013 | 0.029 | 0.67 | **-0.08** | **0.036** | **0.03** |
| Model 2 | **EPA** | **-0.098** | **0.042** | **0.02** | -0.011 | 0.054 | 0.83 | -0.065 | 0.054 | 0.23 | **0.08** | **0.035** | **0.02** | 0.069 | 0.063 | 0.28 | -0.13 | 0.078 | 0.09 |
| | **AA** | **-0.21** | **0.093** | **0.02** | 0.04 | 0.12 | 0.74 | -0.21 | 0.12 | 0.082 | **0.16** | **0.08** | **0.049** | 0.19 | 0.14 | 0.18 | -0.2 | 0.17 | 0.25 |
| | **Interaction** | **-0.045** | **0.019** | **0.02** | -0.0018 | 0.024 | >0.80 | -0.035 | 0.024 | 0.16 | **0.04** | **0.016** | **0.03** | 0.031 | 0.029 | 0.27 | -0.054 | 0.035 | 0.12 |
| Model 3 | **EPA** | **-0.098** | **0.042** | **0.02** | -0.011 | 0.054 | >0.80 | -0.062 | 0.054 | 0.25 | **0.08** | **0.035** | **0.02** | 0.069 | 0.063 | 0.28 | -0.13 | 0.078 | 0.09 |
| | **AA** | **-0.21** | **0.093** | **0.024** | 0.04 | 0.12 | 0.74 | -0.2 | 0.12 | 0.10 | **0.16** | **0.08** | **0.049** | 0.19 | 0.14 | 0.18 | -0.2 | 0.17 | 0.25 |
| | **Interaction** | **-0.045** | **0.019** | **0.02** | -0.0018 | 0.024 | >0.80 | -0.035 | 0.024 | 0.15 | **0.04** | **0.016** | **0.03** | 0.031 | 0.029 | 0.27 | -0.054 | 0.035 | 0.12 |
| Model 4 | **EPA** | **-0.098** | **0.04** | **0.02** | -0.011 | 0.053 | >0.80 | -0.07 | 0.052 | 0.18 | **0.08** | **0.035** | **0.02** | 0.064 | 0.063 | 0.31 | -0.13 | 0.077 | 0.09 |
| | **AA** | **-0.22** | **0.091** | **0.02** | 0.046 | 0.12 | 0.70 | **-0.24** | **0.12** | **0.04** | 0.15 | 0.079 | 0.07 | 0.18 | 0.14 | 0.19 | -0.19 | 0.17 | 0.27 |
| | **Interaction** | **-0.046** | **0.018** | **0.01** | -0.0001 | 0.024 | >0.80 | -0.041 | 0.024 | 0.085 | **0.03** | **0.016** | **0.03** | 0.031 | 0.028 | 0.28 | -0.052 | 0.035 | 0.13 |
| Model 1 | **DHA** | **-0.14** | **0.062** | **0.03** | -0.086 | 0.077 | 0.27 | -0.016 | 0.078 | 0.84 | 0.07 | 0.051 | 0.2 | -0.047 | 0.091 | 0.61 | -0.14 | 0.11 | 0.22 |
| | **AA** | **-0.18** | **0.093** | **0.052** | -0.094 | 0.12 | **0.42** | -0.017 | 0.12 | 0.88 | 0.04 | 0.077 | 0.59 | -0.073 | 0.14 | 0.59 | -0.11 | 0.17 | 0.50 |
| | **Interaction** | **-0.063** | **0.028** | **0.02** | -0.043 | 0.035 | **0.21** | -0.0006 | 0.035 | 0.99 | 0.02 | 0.023 | 0.34 | -0.029 | 0.041 | 0.48 | -0.061 | 0.051 | 0.22 |
| Model 2 | **DHA** | **-0.17** | **0.06** | **0.004** | -0.062 | 0.077 | 0.42 | -0.092 | 0.077 | 0.24 | 0.10 | 0.051 | 0.06 | 0.00074 | 0.091 | 0.99 | -0.15 | 0.11 | 0.18 |
| | **AA** | **-0.24** | **0.09** | **0.007** | -0.05 | 0.12 | 0.67 | -0.17 | 0.12 | 0.15 | 0.11 | 0.076 | 0.15 | 0.026 | 0.14 | 0.85 | -0.15 | 0.17 | 0.38 |
| | **Interaction** | **-0.076** | **0.027** | **0.0045** | -0.029 | 0.034 | 0.40 | -0.039 | 0.034 | 0.26 | 0.04 | 0.023 | 0.096 | -0.003 | 0.041 | 0.94 | -0.065 | 0.0498 | 0.19 |
| Model 3 | **DHA** | **-0.17** | **0.06** | **0.004** | -0.062 | 0.077 | 0.42 | -0.088 | 0.077 | 0.25 | 0.10 | 0.051 | 0.06 | 0.00074 | 0.091 | 0.99 | -0.15 | 0.11 | 0.18 |
| | **AA** | **-0.24** | **0.09** | **0.007** | -0.05 | 0.12 | 0.67 | -0.16 | 0.12 | 0.16 | 0.11 | 0.076 | 0.15 | 0.026 | 0.14 | 0.85 | -0.15 | 0.17 | 0.38 |
| | **Interaction** | **-0.076** | **0.027** | **0.0045** | -0.029 | 0.034 | 0.40 | -0.041 | 0.034 | 0.24 | 0.04 | 0.023 | 0.096 | -0.003 | 0.041 | 0.94 | -0.065 | 0.0498 | 0.19 |
| Model 4 | **DHA** | **-0.13** | **0.058** | **0.02** | -0.047 | 0.076 | 0.54 | -0.038 | 0.075 | 0.62 | 0.09 | 0.051 | 0.087 | 0.042 | 0.09 | 0.64 | -0.13 | 0.11 | 0.23 |
| | **AA** | **-0.2** | **0.088** | **0.02** | -0.018 | 0.12 | >0.80 | -0.13 | 0.111 | 0.26 | 0.09 | 0.077 | 0.23 | 0.082 | 0.14 | 0.55 | -0.12 | 0.17 | 0.47 |
| | **Interaction** | **-0.062** | **0.026** | **0.02** | -0.019 | 0.034 | 0.58 | -0.026 | 0.033 | 0.44 | 0.03 | 0.022 | 0.13 | 0.015 | 0.04 | 0.71 | -0.056 | 0.05 | 0.26 |

Model 1: adjust outcome measurements at exam 1;
Model 2: model 1+ age, sex, race, site;

Model 3: model 2+ selected variables from fatty acid cluster leads, including trans-oleic acid, palmitoleic acid, behenic acid, gadoleic acid, arachidonic acid, myristic acid, stearic acid, using stepwise with BIC as model selection criteria;

Model 4: model 3 + selected variables from smoking, BMI, nonhdl, TG, EGFR, systolic blood pressure, diastolic blood pressure, heart rate, ACEI, ARB, beta-blockers, fasting glucose, diabetes, oral hypoglycemic, diuretics, statin, hypolipidemic, albuminuria, using stepwise with BIC as model selection criteria

**Supplemental Table 3: Model Summary**

Sample size: 1325 (males); 1476 (females); total = 2801

Number of distinct sample moments: 130

*Samples per distinct moment: 22*

Number of distinct parameters to be estimated: 87

*Samples per distinct parameter to be estimated: 32*

Degrees of freedom (130 - 87): 43

Chi-square = 41.96

Cmin/dF= 0.976

Probability level = 0.52

CFI = 1.000

**Supplemental report on results:**

Only five parameters required separate, sex-specific estimates. Three were related to age and LVM or EDV; a third related the change in LVM. Only one parameter estimating PUFA-dependent changes was sex-specific: the direct effect of EPA on Exam 5 EDV.

**Longitudinal changes in outcomes:** Exam 1 and Exam 5 values, expressed as indexed and adjusted to 10 years, for LVM, EDV, and ESV are shown in **Supplemental Table 2** along with the additional calculated estimands, $EF_{calc}$ and $M{:}V_{calc}$. Over 10 years, males had increased in LVM and a decrease in EDV without a decrease in ESV. Hence, $EF_{calc}$ decreased but $M{:}V_{calc}$ did not change. Over the same period females had a decrease in mass, a large decrease in EDV matched with a correspondingly large decrease in ESV which prevented a decline in $EF_{calc}$ or change in $M{:}V_{calc}$.

**Supplemental Table 4**

| | | Male<br>Mean (95% CI) | | Female<br>Mean (95% CI) | |
|---|---|---|---|---|---|
| **LVMpct**[A] | **Exam 1** | 82.5 (81.8, 83.2) | | 86.4 (85.8, 87.0) | |
| | **Exam 5** | 86.9 (86.1, 87.7) | | 84.6 (83.9, 85.3) | |
| | **$\Delta_{E5\text{-}E1}$**[B] | 5.4% (4.6, 6.2)[B] | <0.001 | -2.1% (-2.8, -1.4)[B] | <0.001 |
| **EDVpct**[A] | **Exam 1** | 87.0 (86.1, 87.9) | | 99.3 (98.5, 100.1) | |
| | **Exam 5** | 81.4 (80.3, 82.4) | | 88.2 (87.3, 89.0) | |
| | **$\Delta_{E5\text{-}E1}$** | -5.7 (-6.6, -4.7) | <0.001 | -11.2 (-11.9, -10.4) | <0.001 |
| **ESVpct**[A] | **Exam 1** | 33.6 (33.2, 34.1) | | 35.9 (35.5, 36.3) | |
| | **Exam 5** | 33.1 (32.5, 33.8) | | 31.9 (31.4, 32.4) | |

| | | | | | |
|---|---|---|---|---|---|
| | **Δ$_{E5-E1}$** | -0.5 (-1.1, 0.1) | 0.10 | -4.0 (-4.5, -3.6) | <0.001 |
| **EF%$_{calc}$** | **Exam 1** | 61.4 (61.0, 61.7) | | 63.8 (63.5, 64.1) | |
| | **Exam 5** | 59.3 (58.8, 59.7) | | 63.8 (63.5, 64.2) | |
| | **Δ$_{E5-E1}$** | -2.08 (-2.58, -1.59) | <0.001 | 0.00 (-0.39, 0.38) | >0.80 |
| **M:V$_{calc}$$^{C}$** | **Exam 1** | 0.95 (0.94, 0.96) | | 0.87 (0.86, 0.88) | |
| | **Exam 5** | 0.97 (0.93, 1.01) | | 0.86 (0.82, 0.89) | |
| | **Δ$_{E5-E1}$** | 0.022 (-0.014, 0.059) | 0.24 | -0.013 (-0.047, 0.023) | 0.47 |

$^{A}$ Indexed, Liao *et al*[1].

$^{B}$ Percent is proportional change in Indexed.

$^{C}$ Calculated as (LVM)/(EDV).

**Assessment of the direct, indirect, and total effect of PUFAs:** The pathway model accounts for the multiple ways in which PUFAs can impact both baseline and time-dependent changes in left ventricle structure and function. PUFA levels in healthy Americans who are not taking supplements or ω3-PUFA pharmaceuticals remain stable over at least seven years of observation (22623386), hence it is reasonable to suggest that PUFA levels observed at Exam 1 approximate PUFA levels prior to Exam 1 and through Exam 5. Further, it is also plausible to suggest that EPA and AA have already exerted some effect on baseline ventricular structure and function, and that the state of the LV at Exam 1 impacts the state of the LV at Exam 5. Importantly, our model accounts for the effect of AA and EPA to establish the baseline state of the LV at Exam 1, and accounts for the indirect effect of PUFAs on the observed Exam 5 status by way of their effect on the status at Exam 1. In more practical terms, this means that the LVM observed at Exam 5 is a function of the direct effects of EPA and AA on LVM at Exam 5 combined with their indirect effects on LVM at Exam 1. **In sum:**

***Indirect effects*** of PUFAs on LVM, EDV, and ESV at Exam 5 occur when:

a. There is a significant association of a PUFA with the LV status at Exam 1 *and*

b. There is a significant association of the LV status between Exam 1 and Exam 5.

When both criteria are met, there is strong potential for a significant, indirect effect. Indirect effects represent the PUFA-associated differences from the Exam 1 mean, and consequentially their association with an ultimate difference in Exam 5 because of the association of Exam 1 with Exam 5. The indirect effects are said to be "effects of $PUFA_x$ on Exam $5_{state}$ mediated by Exam $1_{state}$."

***Direct effects*** of PUFAs on Exam 5 occur when:

c. There is a PUFA-associated difference in the state of the LV compared to the Exam 5 mean.

d. Direct effects represent the outcome state at Exam irrespective of its initial status, and are reported in the main text as <u>*Final outcome status*</u>.

***Total effects*** of PUFAs occur when:

e. The cumulative difference in the LV state between Exam 1 and Exam 5 that is attributable to PUFAs ≠ 0.

f. Total effects represent the cumulative difference from Exam 1 to Exam, and are reported in the main text as <u>*Change in outcome*</u>

***Effects are conditional*** when:

g. There is a significant interaction between %AA and %EPA such that the magnitude of the EPA effect depends on the amount of AA present and *vice versa*. Any indirect or direct effect can be conditional if it is dependent on the amount of EPA or AA.

**Indirect effects of AA and EPA on LVM, EDV and ESV at Exam 5:** Indirect effects arising from an association of AA or EPA with LVM, EDV, or EF at Exam 1 are summarized in **Supplemental Table 4** and **Supplemental Figures 1, 3, and 5** for LVM, EDV, and ESV respectively. The figures do not consider biological limits to AA×EPA mixtures. Not all associations had predictive value and were excluded for parsimony; for example, neither AA nor EPA were predictive of LVM at Exam 1, hence there were no indirect effects of either PUFA on any Exam 5 status mediated by Exam 1 LVM. Parameters were identical for males and females by default and freed only when required for model fit. The sole PUFA-dependent example was the effect of EPA on EDV at Exam 5 (Supplemental Table 4) which required separate terms for males and females; neither was significantly different from zero, however they were significantly different from each other and model fit was superior with these sex-specific parameters.

The most influential indirect effect on all Exam 5 metrics was the conditional indirect effect of EPA×AA mediated by Exam 1 EDV **(Supplemental Figures 1, 3, and 5)**. This effect was strong since there was a significant interaction between %AA and %EPA on EDV at Exam 1, which in turn had a strong, independent association with LVM, EDV, and ESV at Exam 5. For example, the indirect effect of EPA on LVM at Exam 5 mediated by EDV at Exam 1 depended on how much AA was present: when AA was low, EPA increased LVM at Exam 5. Conversly, when AA was high, EPA decreased LVM at Exam 5 (**Supplemental Figure 1A-B**). This was also true for the indirect effects of AA on LVM at Exam 5 mediated by EDV at Exam 1 (**Supplemental Figure 1C-D**). Indirect effects of EPA mediated by ESV at Exam 1 were sex-dependent, but not conditional. They had an association with lower LVM at Exam 5 for males, but not females, and a lower ESV at Exam 5 for both sexes. We did not find any effects of AA on Exam 5 status mediated by Exam 1 ESV. Indirect effects of either PUFA mediated by LVM were not found.

**Direct Effects of AA and EPA on LVM, EDV, and ESV at Exam 5:** Conditional direct effects (CDE) as well as total conditional effects (TCE), which are the sum of the indirect and direct effects, are plotted with error estimates but without regard to biological limits to AA×EPA mixtures in **Supplemental Figures 2, 4, and 6** for LVM, EDV, and ESV at Exam 5 respectively. DEs for AA and EPA were present for all outcomes, but conditional only for LVM and ESV. For LVM at Exam 5, higher levels of AA were required for EPA to have a negative association with mass **(Supplemental Figure 2A, B)**. In contrast, higher levels of EPA were required for AA to have a positive association with mass **(Supplemental Figure 2C, D)**.

For EDV at Exam 5 there was no AA×EPA interaction, hence the effect of AA and EPA were independent, not conditional. EPA had a positive association with EDV at Exam 5 among males (p=0.03), but not among females (p=0.40) **(Supplemental Figure 4A, B)**. AA had a positive association with EDV at Exam 5 (p=0.03) irrespective of sex **(Supplemental Figure 4C, D)**.

For ESV at Exam 5, higher levels of AA were required for EPA to have a negative association with mass **(Supplemental Figure 6A, B)**. In contrast, higher levels of EPA were required for AA to have a positive association with mass **(Supplemental Figure 6C, D)**.

**Total conditional effects of PUFAs on LV status at Exam 5:** All total effects were conditional (TCE), representing the cumulative effect of direct and indirect effects. In the case of LVM, the prevalent change independent of PUFAs was an age-dependent decline for both sexes, but more so in females. Hence, a positive TCE counteracts normal aging but a negative effect counteracts it. LVM the CDE was much larger than the CIE, and so the TCE was nearly identical to the CDE. For both males and females, the age-related change was a decline in LVM, and so EPA counteracted this decline only when %AA was low **(Supplemental Figure 2A, B)**. In contrast, AA counteracted the decline only when %EPA was high **(Supplemental Figure 2C, D)**, making for a counter-intuitive interpretation of PUFA-dependent effects.

The total indirect effects on EDV at Exam 5 were much larger relative to the direct effects. This means that the relationship of PUFAs to Exam 1 status contributed a greater share of the total PUFA-dependent changes **(Supplemental Figure 4)**. For EDV, the reference change (*i.e*. change independent of PUFAs) was an age-dependent decline for both sexes, but more so in females. In males with low %AA, the CTE of EPA (combined direct and indirect effects) complemented each other and countereacted the age-dependent decline. However with higher %AA, the direct effect of EPA was nullified by the indirect effect. In females, the CTE of EPA had no cumulative effect on EDV except at the highest %AA, where it accelerated the decline. This was largely due to the small, non-significant direct effect. In contrast to EPA, For both males and females, the direct effect of AA was to counteract age-dependent declines in EDV, however higher %EPA by way of its indirect effect nullified the delayed decline.

The total indirect effects on ESV at Exam 5 were complementary for both sexes and PUFAs **(Supplemental Figure 6)**. For ESV, the reference change was sex-dependent: males had no age-dependent decline, however females had an age-dependent decline of 4%. In males with low %AA, there was nearly no total effect, however in the presence of increasingly higher %AA, EPA was associated with declines in total ESV. In females, this represents a decline beyond the age-dependent decline, in males it is a decline that is otherwise not present.

The indirect of AA was small, hence the TCE was nearly identical to the CDE and nearly identical for both sexes. In the absence of abundant %EPA, AA was associated with greater than reference ESV at Exam 5. Since both males and females had a decline in EDV, this would be expected to produce or exacerbate a decline in ejection fraction. However, with greater %EPA this effect was diminished to zero. In males this could counteract age-dependent declines in EDV, however higher %EPA by way of its indirect effect nullified the delayed decline.

**Combined AA×EPA effects on LV status at Exam 5:** A limitation of the individual presentation of condidtional effects shown in **Supplemental Figures 1-6** is that: 1) there is no restriction to only AA×EPA combinations that occur in vivo; 2) translation to specific AA×EPA combinations is not inuitive; 3) it is not obvious whether at AA×EPA combination the final PUFA effect results in a significant total effect or final Exam 5 Status; finally, it is not clear how the PUFA-dependent effects on LVM, EDV, and ESV affect two other important clinical parameters that can be calculated from these three parameters – ejection fraction (EF) and mass to volume ratio (M:V). **Figures 4-5** summarize the directly estimated LVM, EDV, and ESV using heatmaps and plotting ony the AAxEPA combinations in this cohort having >5th percentile prevalence from Figure 2 with significance as indicated. Further, EF and M:V are provided in **Figures 6 and 7** as calculated posteriorly from estimands. Each figure further provides the reference Exam 1 and Exam 5 values to understand the differences represented by the heatmap color intensity.

**Supplemental Table 5: Model Parameter Estimates**

| | | Outcome | Shared[A] | p-val | Male[B] | p-val | Female[C] | p-val |
|---|---|---|---|---|---|---|---|---|
| **2-step indirect** | | | | | | | | |
| **1. Exogenous predictors on Exam 1** | | | | | | | | |
| AA[D] | ---> | E1 LVM | . | . | . | . | . | . |
| EPA[D] | ---> | E1 LVM | . | . | . | . | . | . |
| AAxEPA[D] | ---> | E1 LVM | . | . | . | . | . | . |
| **Age** | **--->** | **E1 LVM** | . | . | -0.00021 ± 0.00018 | 0.25 | **0.00041 ±0.00017** | **0.01** |
| **AA** | **--->** | **E1 EDV** | **-0.80 ±0.27** | **0.004** | . | . | . | . |

| | | | | | | | | |
|---|---|---|---|---|---|---|---|---|
| EPA$^D$ | ---> | E1 EDV | . | . | . | . | . | . |
| **AAxEPA** | **--->** | **E1 EDV** | **-0.34 ± 0.15** | **0.02** | . | . | . | . |
| **Age** | **--->** | **E1 EDV** | **-0.35 ±0.03** | **<0.001** | . | . | . | . |
| AA$^D$ | ---> | E1 ESV | . | . | . | . | . | . |
| **EPA** | **--->** | **E1 ESV** | **-0.24 ± 0.10** | **0.01** | . | . | . | . |
| AAxEPA$^D$ | ---> | E1 ESV | . | . | . | . | . | . |
| **Age** | **--->** | **E1 ESV** | **-0.14 ± 0.02** | **<0.001** | . | . | . | . |
| **2. Endogenous, Exam 1 on Exam 5** | | | | | | | | |
| **LVM** | **--->** | **LVM** | . | . | **0.60 ± 0.03** | **<0.001** | **0.54 ± 0.03** | **<0.001** |
| **EDV** | **--->** | **LVM** | **0.0005 ±0.00008** | **<0.001** | . | . | . | . |
| ESV | ---> | LVM | 0.0003 ±0.0002 | 0.07 | . | . | . | . |
| **LVM** | **--->** | **EDV** | . | . | **17 ± 4.6** | **<0.001** | **12 ± 4.6** | **0.01** |
| **EDV** | **--->** | **EDV** | **0.49 ± 0.03** | **<0.001** | . | . | . | . |
| **ESV** | **--->** | **EDV** | **0.22 ± 0.05** | **<0.001** | . | . | . | . |
| LVM | ---> | ESV | . | . | . | . | . | . |
| **EDV** | **--->** | **ESV** | **0.15 ± 0.02** | **<0.001** | . | . | . | . |
| **ESV** | **--->** | **ESV** | **0.43 ± 0.03** | **<0.001** | . | . | . | . |
| **Direct – Exogenous on Exam 5** | | | | | | | | |
| **AA** | **--->** | **LVM** | **0.0033 ± 0.0011** | **0.002** | . | . | . | . |
| EPA$^D$ | ---> | LVM | . | . | . | . | . | . |
| **AAxEPA** | **--->** | **LVM** | **-0.0024 ± 0.0008** | **0.003** | . | . | . | . |
| **Age** | **--->** | **LVM** | . | . | **-0.00049 ± 0.00017** | **0.004** | -0.00003 ± 0.00017 | 0.85 |
| **AA** | **--->** | **EDV** | **0.60 ± 0.25** | **0.02** | . | . | . | . |
| EPA | ---> | EDV | . | . | 0.55 ± 0.36 | 0.13 | -0.18 ± 0.30 | 0.54 |
| AAxEPA$^D$ | ---> | EDV | . | . | . | . | . | . |
| **Age** | **--->** | **EDV** | . | . | **-0.17 ± 0.04** | **<0.001** | **-0.29 ± 0.04** | **<0.001** |
| **AA** | **--->** | **ESV** | **0.60 ± 0.25** | **0.001** | . | . | . | . |
| EPA | ---> | ESV | -0.20 ± 0.15 | 0.18 | . | . | . | . |
| **AAxEPA** | **--->** | **ESV** | **-0.20 ± 0.10** | **0.047** | . | . | . | . |
| **Age** | **--->** | **ESV** | **-0.044 ± 0.018** | **0.01** | . | . | . | . |

Significant Parameter estimates in **bold**, $p<0.05$.

[A] Parameter is shared among both male and female participants
[B] Male-specific parameter, using sex-specific parameters optimizes model fit.
[C] Female-specific parameter, using sex-specific parameters optimizes model fit.
[D] Parameter was dropped from model for parsimony.

### Supplemental Table 6: Model Means and intercepts

| | Shared[A] | | Male[B] | | Female[C] | |
|---|---|---|---|---|---|---|
| **Exogenous means** | | | | | | |
| **AA** | . | . | **-0.141 ±0.029** | <0.001 | 0.037 ±0.027 | 0.18 |
| **EPA** | . | . | 0.024 ±0.028 | 0.38 | **0.143 ±0.028** | **<0.001** |
| AAxEPA | 0.043 ±0.024 | 0.08 | . | . | . | . |
| **Age** | **59.5 ±0.2** | **<0.001** | . | . | . | . |
| **Endogenous intercepts** | | | | | | |
| ***Exam 1*** | | | | | | |
| **E1 LVM** | **.** | **.** | **1.93 ±0.01** | **<0.001** | **1.91 ±0.01** | **<0.001** |
| **E1 EDV** | **.** | **.** | **107.9 ±1.9** | **<0.001** | **120.4 ±1.9** | **<0.001** |
| **E1 EF** | **.** | **.** | **61.3 ±0.2** | **<0.001** | **63.7 ±0.1** | **<0.001** |
| ***Exam 5*** | | | | | | |
| **E5 LVM** | **.** | **.** | **0.8 ±0.04** | **<0.001** | **0.75 ±0.04** | **<0.001** |
| **E5 EDV** | . | . | 3.9 ±13 | 0.77 | **40.8 ±11.1** | **<0.001** |
| **E5 EF** | **.** | **.** | **41 ±4** | **<0.001** | **46 ±4** | **<0.001** |

Significant Parameter estimates in **bold**, p<0.05.

[A] Parameter is shared among both male and female participants

[B] Male-specific parameter, parameter is different between sexes.

[C] Female-specific parameter, parameter is different between sexes.

**Supplemental Table 7: Model Covariances**

| Covariance | | | Shared[A] | | Male[B] | | Female[C] | |
|---|---|---|---|---|---|---|---|---|
| **Exogenous** | | | **Estimate, SE** | **p-val** | **Estimate, SE** | **p-val** | **Estimate, SE** | **p-val** |
| | **EPA** | **Age** | . | . | 0.40 ±0.24 | 0.10 | **1.33 ±0.25** | **<0.001** |
| | Age | AA | . | . | -0.49 ±0.27 | 0.07 | **0.57 ±0.25** | **0.02** |
| | Age | AAxEPA[D] | . | . | . | . | . | . |
| | **EPA** | **AA** | **0.043 ±0.022** | **0.05** | . | . | . | . |
| | **AA** | **AAxEPA** | . | . | **-0.14 ±0.04** | **<0.001** | -0.01 ±0.03 | >0.80 |
| | **EPA** | **AAxEPA** | . | . | **-0.74 ±0.04** | **<0.001** | **-0.46 ±0.04** | **<0.001** |
| **Exam 1** | | | | | | | | |
| | **E1 LVM** | **E1 EDV** | **.** | **.** | **0.54 ±0.03** | **<0.001** | **0.5 ±0.03** | **<0.001** |
| | E1 EDV | E1 EF | . | . | 1.97 ±2.55 | 0.44 | 2.58 ±2.11 | 0.22 |
| | **E1 LVM** | **E1 EF** | **.** | **.** | **-0.039 ±0.011** | **<0.001** | -0.009 ±0.009 | 0.32 |
| **Exam 5** | | | | | | | | |
| | **E5 LVM** | **E5 EDV** | **.** | **.** | **0.36 ±0.03** | **<0.001** | **0.29 ±0.02** | **<0.001** |
| | **E5 EDV** | **E5 EF** | **.** | **.** | **-12.3 ±3.1** | **<0.001** | 2.4 ±2.3 | 0.29 |
| | E5 LVM | E5 EF | . | . | -0.018 ±0.012 | 0.11 | 0.011 ±0.01 | 0.28 |

Significant Parameter estimates in **bold**, p<0.05.

[A] Parameter is shared among both male and female participants

[B] Male-specific parameter, parameter is different between sexes.

[C] Female-specific parameter, parameter is different between sexes.

[D] Parameter was dropped from model for parsimony.

### Supplemental Table 8: Model Fit Indices – Minimal Discrepancy

| Model | Number of Parameters | $X^2_{min}$ | dF | P-val | $X^2_{min}/dF$[a] |
|---|---|---|---|---|---|
| **Final Model** | 86 | 42.2 | 44 | 0.55 | 0.96 |
| **Saturated model** | 130 | 0 | 0 | | |
| **Independence model** | 40 | 9604 | 90 | 0 | 106.71 |

(a) recommended <2[2]

### Supplemental Table 9: Model Fit Indices – Minimum Value of Discrepancy

| Model | $F_{MIN}$ | F0 | $LL_{90\%}$ | $UL_{90\%}$ |
|---|---|---|---|---|
| **Final Model** | 0.015 | 0.000 | 0.000 | 0.006 |
| **Saturated model** | 0 | 0 | 0 | 0 |
| **Independence model** | 3.43 | 3.40 | 3.29 | 3.52 |

### Supplemental Table 10: Model Fit Indices – Hoelter's N

| Model | Hoelter p=0.05 | Hoelter p=0.01 |
|---|---|---|
| **Final Model** | 4011 | 4556 |
| **Independence model** | 34 | 38 |

### Supplemental Table 11: Model Fit Indices – Root Mean Square Error of Approximation

| Model | RMSEA | $LL_{90\%}$ | $UL_{90\%}$ |
|---|---|---|---|
| **Final Model** | 0.000 | 0.000 | 0.012 |
| **Independence model** | 0.194 | 0.191 | 0.198 |

### Supplemental Table 12: Model Fit Indices – Information Criteria

| Model | AIC | BCC |
|---|---|---|
| **Final Model** | 214.2 | 215.6 |
| **Saturated model** | 260.0 | 262.0 |
| **Independence model** | 9684 | 9685 |

**Supplemental Figure 1: Detailed Summary of Indirect effects of AA and EPA on Exam 5 LVM*10-year adjusted***

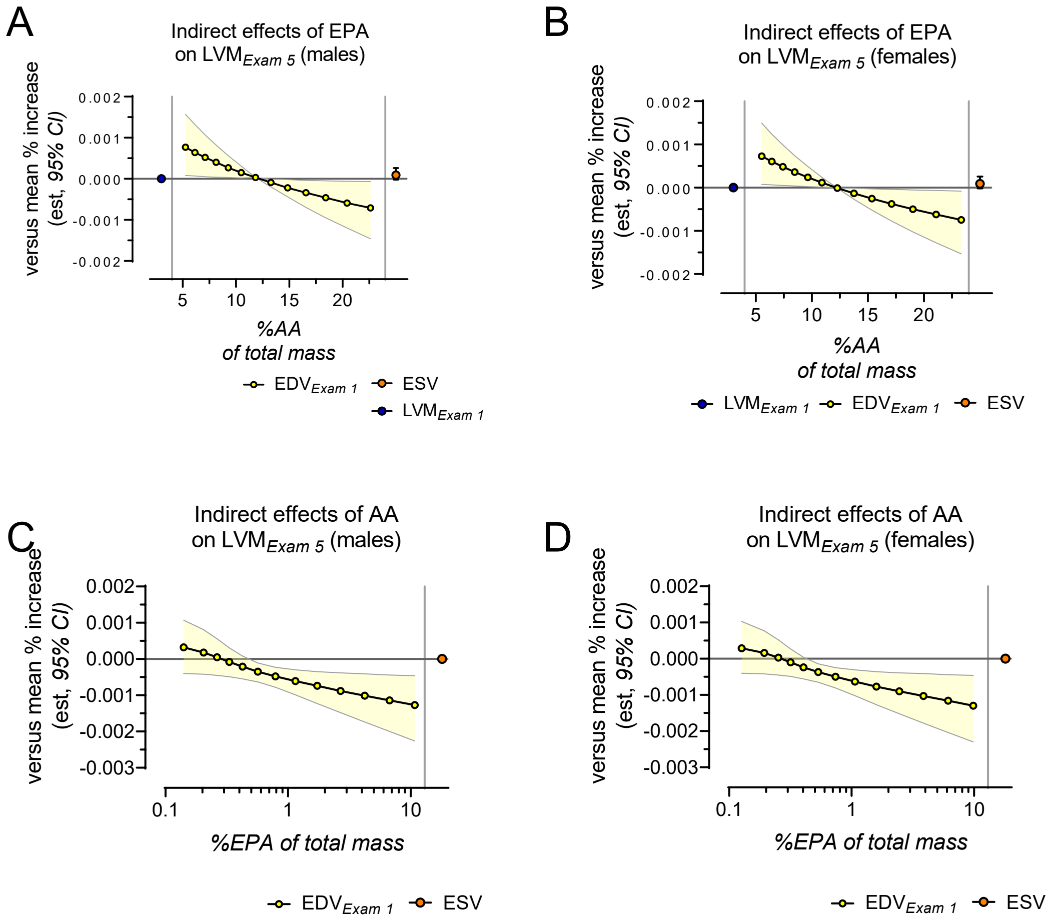

*Edited 2/9/2022 – needs touching up*

*Conditional indirect effects of AA and EPA on Left Ventricular Mass mediated by Exam 1 EDV, and ESV* – The indirect effects of EPA on Exam 5 LVM status in males **(A)** and females **(B)** mediated by Exam 1 status are shown above; the indirect effects of AA on Exam 5 LVM status in males **(C)** and females **(D)** mediated by Exam 1 status are shown below. The most prominent indirect effect of EPA on Exam 5 LVM was mediated by Exam 1 EDV (yellow) and was conditional on %AA among both males and females. In the presence of low %AA, EPA had a positive indirect effect on LVM by means of Exam 1 EDV, but with high %AA, EPA had a negative indirect effect. AA had a negative indirect effect on LVM by means of Exam 1 EDV, but only when %EPA was high.

*Note:* a single point represents non-conditional indirect effects, not depend on the other PUFA. Where no symbol is present, the term was not needed, and eliminated for parsimony.

**Supplemental Figure 2: Summary of Direct and Cumulative Indirect effects on Exam 5 LVM*10-year adjusted***

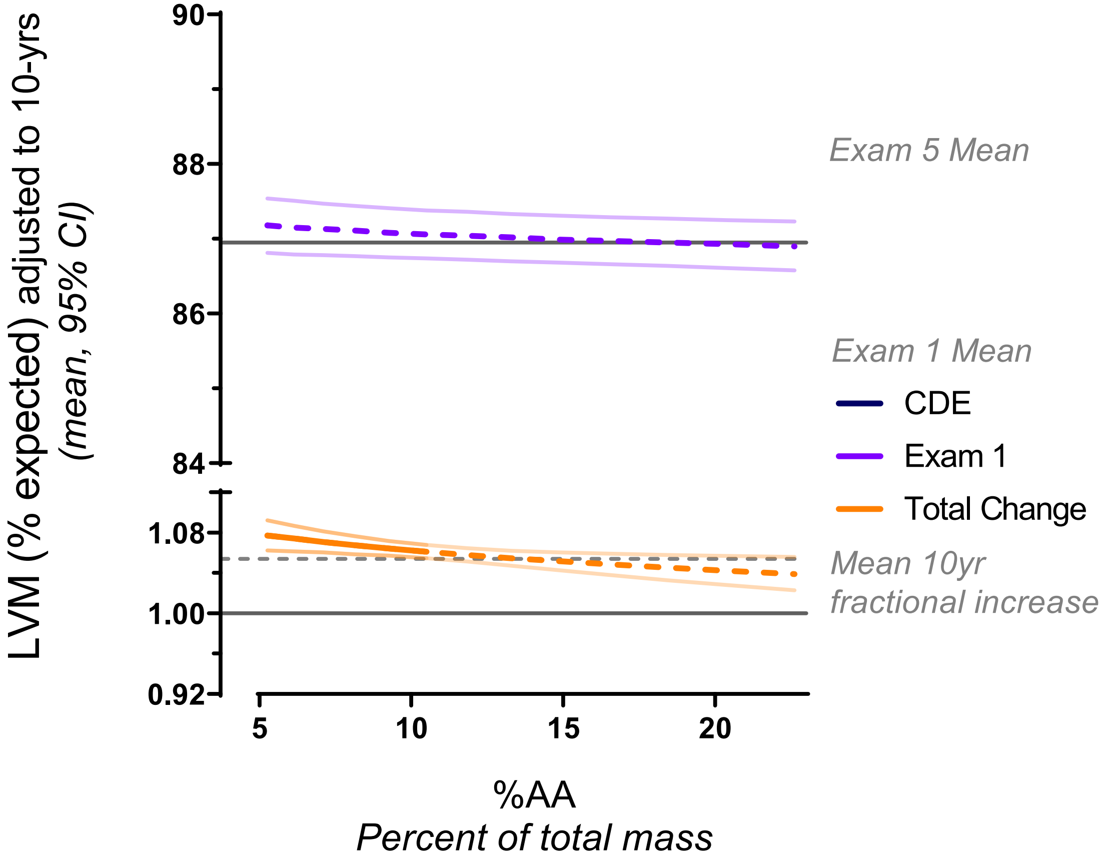

*Conditional direct and total effects of AA and EPA on Left Ventricular Mass* – The conditional direct effect (CDE) of EPA and the total conditional effect (TCE) of EPA, represented as the fractional change from Exam 1, on Exam 5 LVM*10-yr adjusted* are shown in males (A) and females (B) respectively. A positive direct effect of EPA on Exam 5 LVM*10-yr adjusted* was present with low %AA, however this effect declined with elevated AA and disappeared when %AA was ~12% in males and ~10% in females; with higher %AA it tended to be negative. The means that EPA was directly associated with greater than average Exam 5 LVM*10-yr adjusted* when %AA was low. The TCE is the sum of direct and the indirect effects from Supplemental Figure 1; since the direct effects were large compared to the indirect effect, the TCE followed the general pattern of the direct effects irrespective of the sex-dependent changes in LVM.

The CDE of AA on Exam 5 LVM*10-yr adjusted* and the TCE of AA, represented as the fractional change from Exam 1, are shown in males (C) and females (D) respectively. The direct effect of AA on Exam 5 LVM*10-yr adjusted* was highly conditioned on EPA. When %EPA was <~0.4%, AA is associated with smaller than average Exam 5 LVM*10-yr adjusted*, however with higher %EPA. The effect disappeared at higher %EPA until at ~1%, where the effect of AA was associated with greater than average Exam 5 $LVM_{10\text{-}yr\ adjusted}$ in both males and females. The total effect of AA resembled the direct effect of AA.

**Supplemental Figure 3: Detailed Summary of Indirect effects of AA and EPA on Exam 5 EDV$_{\textit{10-year adjusted}}$**

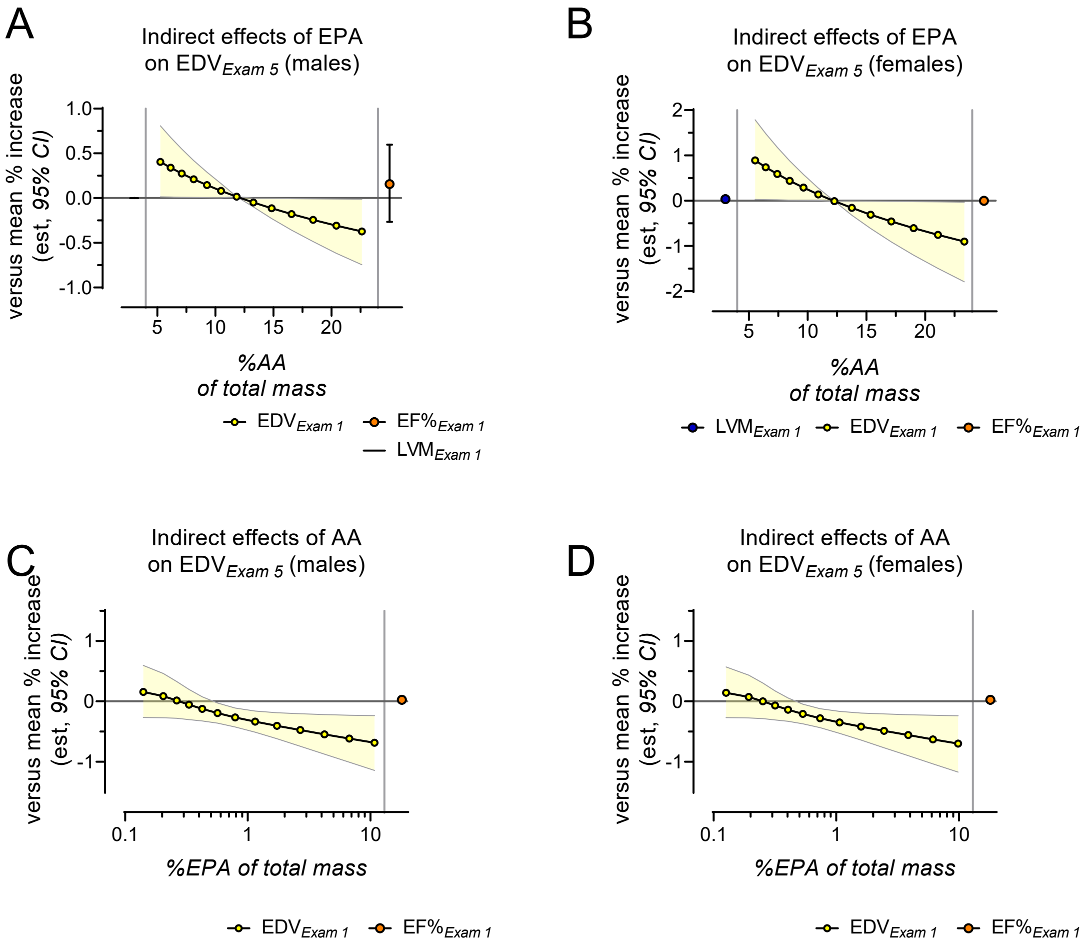

*Conditional indirect effects of AA and EPA on End Diastolic Volume mediated by Exam 1 LVM, EDV, and EF%* – The indirect effects of EPA on Exam 5 EDV$_{\textit{10-yr adjusted}}$ mediated by Exam 1 LVM, EDV, and EF% are shown in males (A) and females (B) respectively. There was no indirect effect of EPA on Exam 5 EDV$_{\textit{10-yr adjusted}}$ mediated by Exam 1 LVM in males or females, nor was there an indirect effect of EPA mediated by Exam 1 EF% on Exam 5 EDV$_{\textit{10-yr adjusted}}$. The indirect effect of EPA mediated by Exam 1 EDV on Exam 5 EDV$_{\textit{10-yr adjusted}}$ was conditional on %AA: it was positive when %AA was low, meaning that under these it is associated with larger than average Exam 5 EDV$_{\textit{10-yr adjusted}}$, however at higher %AA, it was associated with smaller than average Exam 5 EDV$_{\textit{10-yr adjusted}}$.

The indirect effects of AA on Exam 5 EDV$_{\textit{10-yr adjusted}}$ mediated by Exam 1 LVM, EDV, and EF% are shown in males (C) and females (D) respectively. There was no indirect effect of AA on Exam 5 LVM$_{\textit{10-yr adjusted}}$ mediated by Exam 1 LVM or EF%. The indirect effect of EPA on Exam 5 LVM$_{\textit{10-yr adjusted}}$ mediated by Exam 1 EDV was conditional on %EPA: the indirect effect of AA was neutral when

%EPA was low, meaning that under these conditions AA is not associated with time-dependent changes in LVM that are different than average, however at higher %EPA, AA was associated with smaller than average time-dependent changes in Exam 5 $LVM_{10\text{-}yr\ adjusted}$.

*Note:* a single point represents indirect effects that are not conditional on the other PUFA. Where a symbol is absent, the term was eliminated for parsimony.

**Supplemental Figure 4: Summary of Direct and Indirect effects on Exam 5 EDV*10-year adjusted***

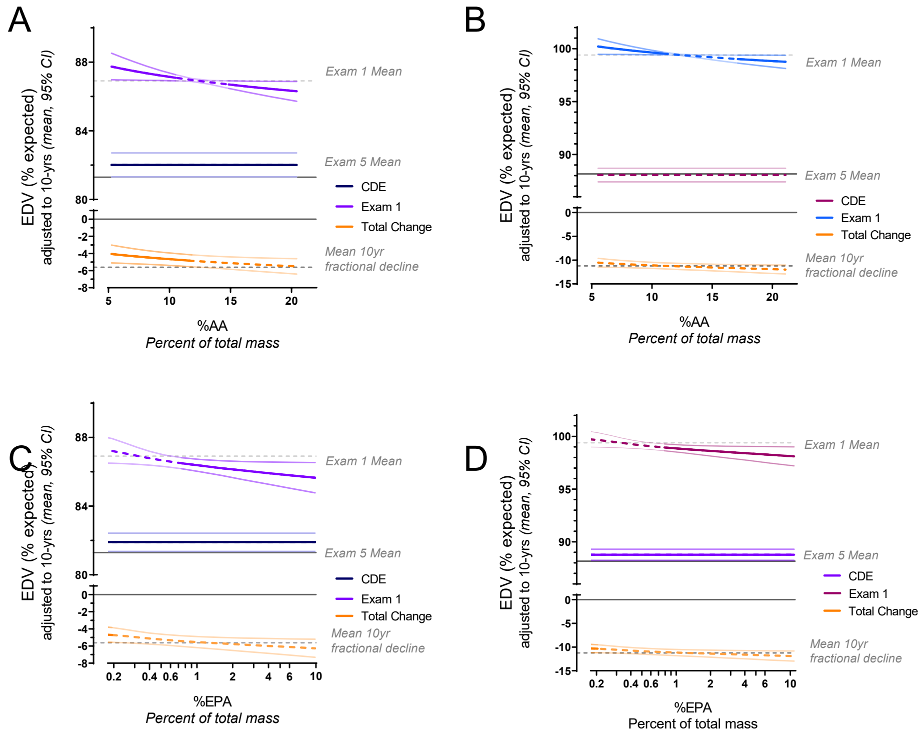

*Conditional direct and total effects of AA and EPA on End Diastolic Volume* – The conditional direct effect (CDE) of EPA and the total conditional effect (TCE) of EPA, represented as the fractional change from Exam 1, on Exam 5 EDV*10-yr adjusted* are shown in males (A) and females (B) respectively. A positive direct effect of EPA on Exam 5 EDV*10-yr adjusted* was not conditional in males, meaning that EPA counteracted the decline in EDV irrespective of %AA. This effect was not present in females. The TCE is the sum of direct and the indirect effects from Supplemental Figure 3; due to the indirect effects of EPA mediated by Exam 1 EDV, the effect of EPA to counteract time-dependent declines in EDV was present only among males with low %AA, and was not present in females at any %AA.

The CDE and the TCE, represented as the fractional change from Exam 1, of AA on Exam 5 EDV*10-yr adjusted* are shown in males (C) and females (D) respectively. The direct effect of AA on Exam 5 EDV*10-yr adjusted* was not dependent on EPA for either sex, meaning that AA counteracted the decline in EDV irrespective of %EPA. The TCE is the sum of direct and the indirect effects of AA from Supplemental Figure 3; due to the indirect effects of AA mediated by Exam 1 EDV, the effect of AA to counteract time-dependent declines in EDV was present only among participants with very low %AA, irrespective of sex.

**Supplemental Figure 5: Detailed Summary of Indirect effects of AA and EPA on Exam 5 EF%*10-year adjusted***

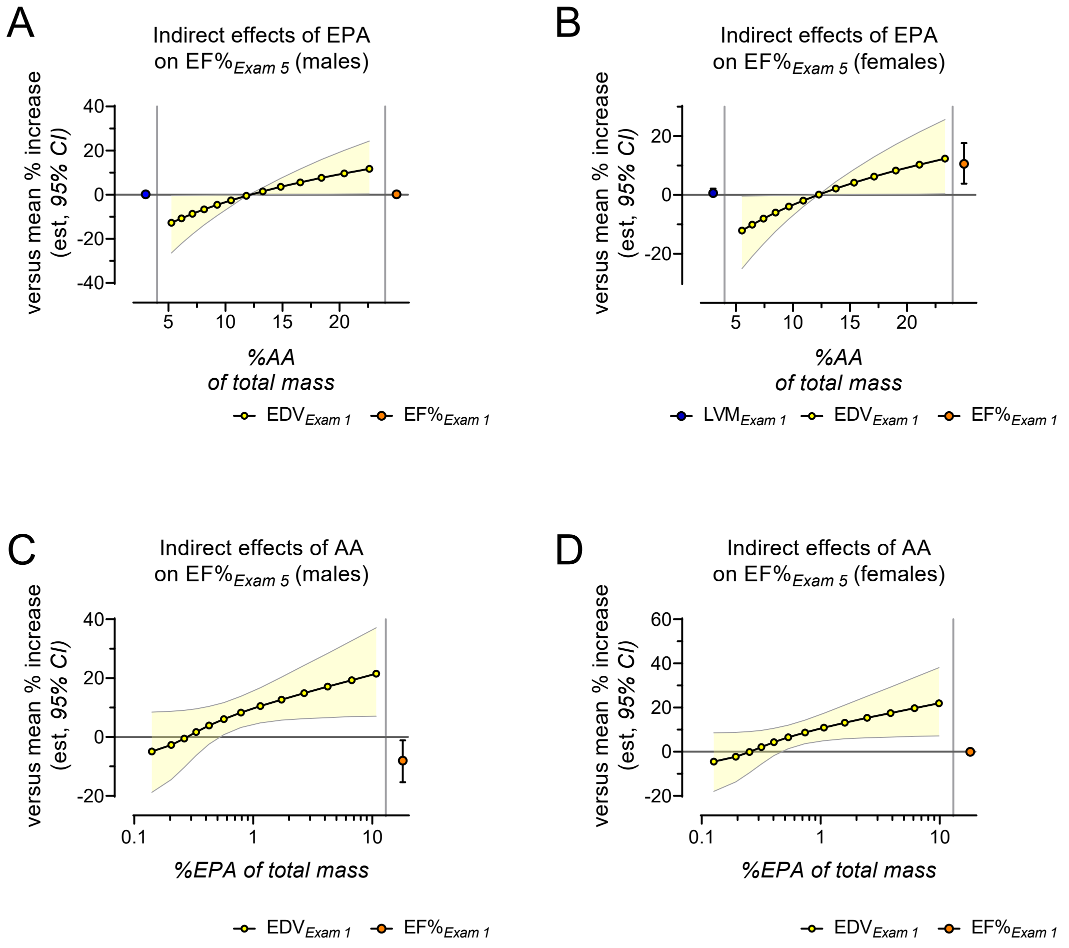

*Conditional indirect effects of AA and EPA on Ejection Fraction mediated by Exam 1 LVM, EDV, and EF%* – The indirect effects of EPA on Exam 5 EF%*10-yr adjusted* mediated by Exam 1 LVM, EDV, and EF% are shown in males (A) and females (B) respectively. There was no indirect effect of EPA on Exam 5 EDV*10-yr adjusted* mediated by Exam 1 LVM in males or females. There was a positive indirect effect of EPA mediated by Exam 1 EF% in females but not in males. The indirect effect of EPA mediated by Exam 1 EDV on Exam 5 EDV*10-yr adjusted* was conditional on %AA: it was negative when %AA was low, meaning that under these conditions it is associated with smaller than average Exam 5 EF%*10-yr adjusted*, however at higher %AA, it was associated with larger than average Exam 5 EF%*10-yr adjusted*.

The indirect effects of AA on Exam 5 EDV*10-yr adjusted* mediated by Exam 1 LVM, EDV, and EF% are shown in males (C) and females (D) respectively. There was no indirect effect of AA mediated by Exam 1 LVM. There was a negative indirect effect mediated by EF% in males, but not in females. The indirect effect of EPA mediated by Exam 1 EDV on Exam 5 EF%*10-yr adjusted* was conditional on %EPA: the indirect effect of AA was neutral when %EPA was low, meaning that under these conditions AA is not associated with

Exam 5 $EF\%_{\textit{10-yr adjusted}}$ that are different than average, however at higher %EPA, AA was associated with larger than average Exam 5 $EF\%_{\textit{10-yr adjusted}}$, irrespective of sex.

*Note:* a single point represents indirect effects that are not conditional on the other PUFA. Where a symbol is absent, the term was eliminated for parsimony.

**Supplemental Figure 6: Summary of Direct and Indirect effects on Exam 5 EF%*10-year adjusted***

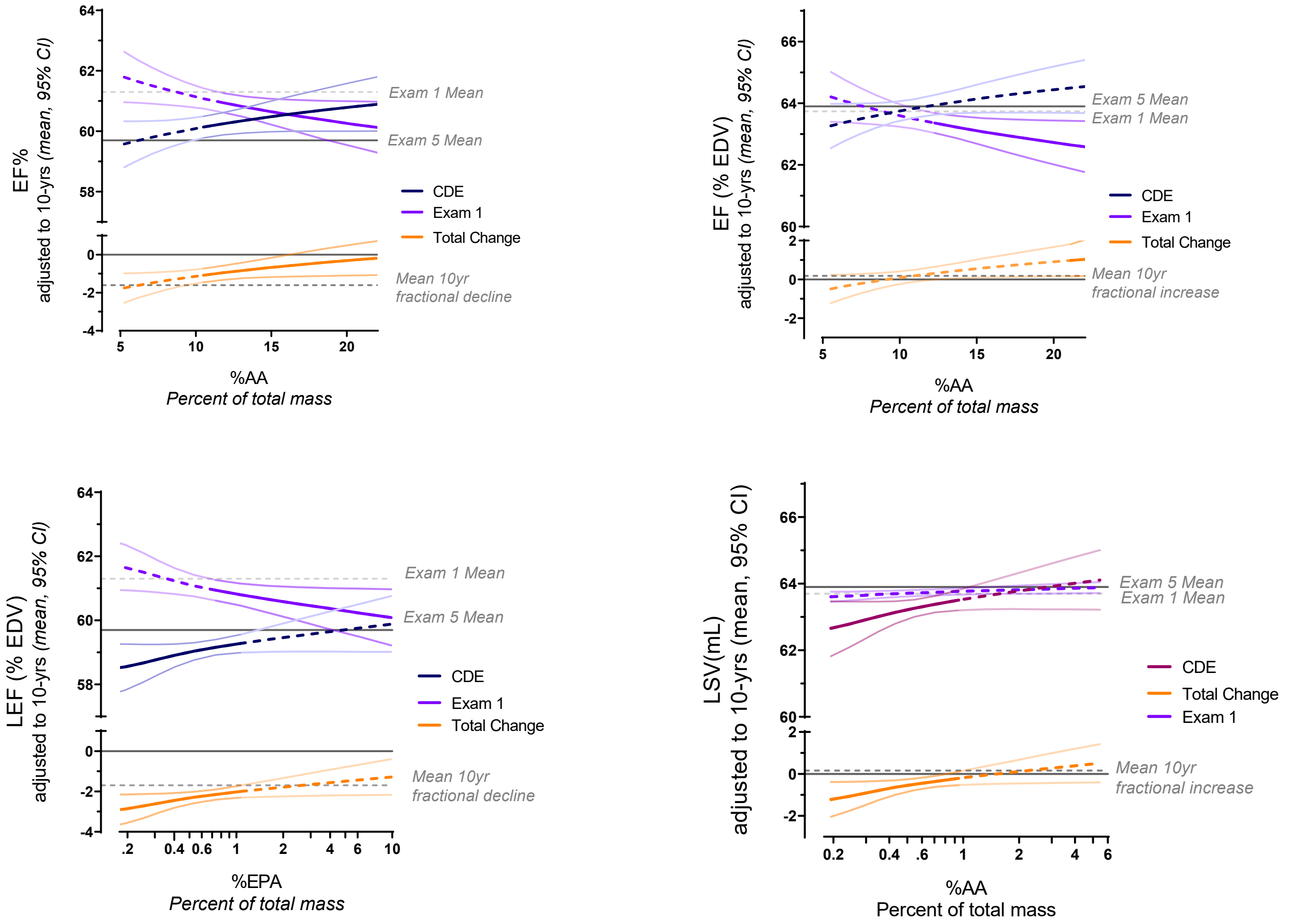

*Conditional direct and total effects of AA and EPA on Ejection Fraction* – The conditional direct effect (CDE) of EPA and the total conditional effect (TCE) of EPA, represented as the fractional change from Exam 1, on Exam 5 EF%*10-yr adjusted* are shown in males (A) and females (B) respectively. In males, the direct effect of EPA on Exam 5 EF%*10-yr adjusted* was conditional in males, meaning that EPA counteracted the decline in EF% among males, but only when %AA was high. The conditionality and directionality of the CDE of EPA was identical in females, but it was not significant in part because there were no significant time-dependent declines in EF% among females. The TCE is the sum of direct and the indirect effects from Supplemental Figure 5: among males, the effect of EPA to counteract time-dependent declines in EF% was significant when %AA was greater than ~11%; among females, the effect of EPA was only significant when %AA was very high, > ~21%.

The CDE and the TCE, represented as the fractional change from Exam 1, of AA on Exam 5 $EF\%_{10\text{-}yr\ adjusted}$ are shown in males (C) and females (D) respectively. The CDE of AA on Exam 5 $EF\%_{10\text{-}yr\ adjusted}$ was identical for both sexes. In the absence of EPA, AA was associated with declines in EF%, notably among males where the time-dependent decline was exacerbated. The TCE is the sum of direct and the indirect effects of AA from Supplemental Figure 3; when EPA was low, AA was associated with smaller than average Exam 5 $EF\%_{10\text{-}yr\ adjusted}$, irrespective of sex. However, with %EPA > ~1%, AA was not associated with Exam 5 $EF\%_{10\text{-}yr\ adjusted}$ different than average, irrespective of sex.

refs

1. Liao, Y. C., R.S.; Durazo-Arvizu, R.; Mensah, G.A.; Ghali, J.K., Prediction of Mortality Risk by Different Methods of Indexation for Left Ventricular Mass. *Journal of the American College of Cardiology* **1997,** *29* (3), 641-647.
2. Byrne, B. M., *A primer of LISREL : basic applications and programming for confirmatory factor analytic models*. Springer-Verlag: New York, 1989; p xii, 184 p.
